# Challenges in Quantifying Upper Extremity Dysmetria in the Scale for the Assessment and Rating of Ataxia (SARA)

**DOI:** 10.64898/2026.11.02.26367096

**Authors:** Defne Apaydin, Jeremy D. Schmahmann

## Abstract

**Background:** Assessments of upper extremity control in ataxia are susceptible to measurement error and inherent variability.

**Objectives:** Quantify trial-to-trial variability in the Scale for the Assessment and Rating of Ataxia (SARA) nose-to-finger (NTF) and finger chase (FC) tasks, and test agreement between scores derived from live assessment and retrospective video review.

**Methods:** 17 patients with cerebellar ataxia were video-recorded performing tasks behind a transparent, calibrated 1-cm grid. NTF tremor amplitude and FC undershoot/overshoot distance derived from live scores were compared with measurements from video review.

**Results:** NTF tremor amplitude and FC under/overshoot varied between consecutive trials. Patients received 2.4 ± 0.7 (NTF) and 2.6 ± 0.8 (FC) different SARA scores for limb excursions within one session. Live and video score match was NTF: 64.7% left, 70.6% right; FC: 64.7% left, 41.2% right. Live scoring often overestimated impairment.

**Conclusions:** Trial-to-trial variability and live/video discordance limit the precision of SARA’s upper-limb tasks.

## Introduction

The cerebellar motor syndrome is characterized by temporospatial errors^1^ manifesting as dysmetria, overshoot (hypermetria) and undershoot (hypometria)^2^. Objective assessment of upper-limb function in cerebellar ataxia is challenging because variability is an integral characteristic of dysmetria^3^. The Scale for the Assessment and Rating of Ataxia (SARA)^4^ includes nose-to-finger (NTF) and finger chase (FC) tasks that require visual estimation of dysmetria and tremor amplitude in real time across consecutive trials. The risk of measurement error from intra- and inter-rater variability and biological variability is relevant in considering primary outcome measures in clinical trials. Here, we examined variability of patient performance in the NTF and FC tasks in consecutive excursions in the same testing session, and the agreement between an examiner’s scores during live, in-person assessment compared to subsequent review of video recordings.

## Methods

### Study Population

We recruited 17 patients from the 2026 National Ataxia Foundation Annual Ataxia Conference (N = 13) and the Massachusetts General Hospital Ataxia Center (N = 4).

### Clinical Outcome Assessments (COAs)

*SARA* assesses ataxia severity across 8 tasks: gait, stance, sitting, speech, finger chase, nose-to-finger, fast alternating hand movements, and heel-shin slide, with appendicular items averaged across sides and a maximum score of 40.

The *Friedreich’s Ataxia Rating Scale Functional Staging* (FARS-Func) measures functional status defined by walking ability. Although gait and upper extremity control are not necessarily similarly affected in ataxia, upper extremity control on a computer mouse task correlates with total ataxia score^5^. We therefore used FARS-Func to classify ataxia into mild: normal or no walking disability (Stages < 2.0); moderate: minimal disability, use of a cane to walk or holding onto walls or furniture (Stages ≥ 2.0 and < 4.0); and severe: requires a walker or is no longer able to walk, requires a wheelchair (Stages ≥ 4.0).

### Procedures

SARA examinations (DA; SARA-certification; dzne.de/CERT^6^) were videotaped at a 45-degree angle that visualized both the examiner’s and patient’s arms. A transparent screen marked with a 1 × 1 cm grid was placed between the examiner and patient to quantify dysmetria amplitude (“tremor”, in SARA terminology) and undershoot/overshoot distance. The testing apparatus is shown in Supplementary Figure S1. Patients touched the examiner’s finger on the screen (NTF) and followed the examiner’s finger keeping their finger close to the screen without touching it (FC). SARA scores were derived from the live assessment according to SARA instructions. Videos were then reviewed on a 24-inch computer monitor by both investigators, counting the number of 1-cm squares between the target (examiner’s finger) and the patient’s finger. We derived a SARA score for each excursion on both the NTF and FC task to determine how many different SARA scores resulted from a single session. We then averaged the 3^rd^, 4^th^, and 5^th^ excursions according to SARA guidelines, and all FC excursions, to *derive a single SARA score for the test session* and compared the degree to which the two methods matched the live score.

### Data Analysis

For each patient and hand, within-patient trial-to-trial variability was quantified from tremor amplitude and dysmetria distance (cm) as the mean, standard deviation (SD), and range across trials, and the mean and SD of absolute change between consecutive trials. Each trial’s amplitude was converted to an item-level SARA score (0–3) using the scale’s cm thresholds (NTF: 1 = <2 cm, 2 = 2–5 cm, 3 = >5 cm; FC: 1 = <5 cm, 2 = 5–15 cm, 3 = >15 cm). The same statistics were computed on these scores. Patient-level values were summarized as mean and SD. Live and video-derived scores were compared per patient and hand and summarized as percent agreement. Differences across severity groups were tested with Kruskal-Wallis tests, followed by Dunn’s post hoc tests with Holm correction for the three pairwise comparisons. Analyses were performed with R 4.6.1.

## Results

There were 17 patients (12 females; 50.82 ± 17.36 years; Tables 1A–1D); 5 with spinocerebellar ataxia type 3 (SCA3), 3 SCA2, 2 SCA1, 2 SCA6, 2 SCA27B, and 1 each SCA7, SCA8, and autoimmune-mediated ataxia. Using FARS-Func, 5 were classified mild, 8 moderate, and 4 severe (Tables 1C–1D). Mean live SARA scores were 1.26 ± 0.79 (NTF) and 1.12 ± 0.57 (FC).

### SARA Nose-to-Finger Trial-to-Trial Variability

Trial-level variability in NTF and FC tasks was examined across the 17 patients. For NTF, mean tremor amplitude of all trials was 1.50 ± 1.80 cm (left hand) and 1.24 ± 1.43 cm (right) (Table 1A). For the right hand, the severe group (3.20 ± 1.78 cm) had higher tremor amplitudes than both mild (0.47 ± 0.39 cm; *p*_Holm_ = .017) and moderate (0.74 ± 0.57 cm; *p*_Holm_ = .032; Table 1C; Supplementary Table S1A). Patients deviated from their mean amplitude by 0.99 ± 0.87 cm (left) and 0.81 ± 0.57 cm (right) (Table 1A); in the right hand, SD was higher in the severe group (1.64 ± 0.15 cm) than in the moderate group (0.51 ± 0.34 cm; *p*_Holm_ = .009; Table 1C; Supplementary Table S1A). Tremor amplitudes across all trials ranged by 2.82 ± 2.53 cm (left) and 2.41 ± 1.66 cm (right), and the range in the right hand was larger in the severe group (4.50 ± 0.58 cm) than the moderate (1.62 ± 1.19 cm; *p*_Holm_ = .014; Table 1C; Supplementary Table S1A). Mean tremor amplitude changed by 1.10 ± 0.99 cm (left) and 0.86 ± 0.79 cm (right) between consecutive trials. Trial-to-trial variability was higher in the severe group than the mild group in both hands (left: 2.19 ± 1.40 vs 0.70 ± 0.77 cm, *p*_Holm_ = .046; right: 2.03 ± 0.66 vs 0.42 ± 0.31 cm, *p*_Holm_ = .017) and than the moderate group in the right hand (2.03 ± 0.66 vs 0.56 ± 0.39 cm; *p*_Holm_ = .019; Table 1C, Figure 1A; Supplementary Table S1A).

**Figure 1.**
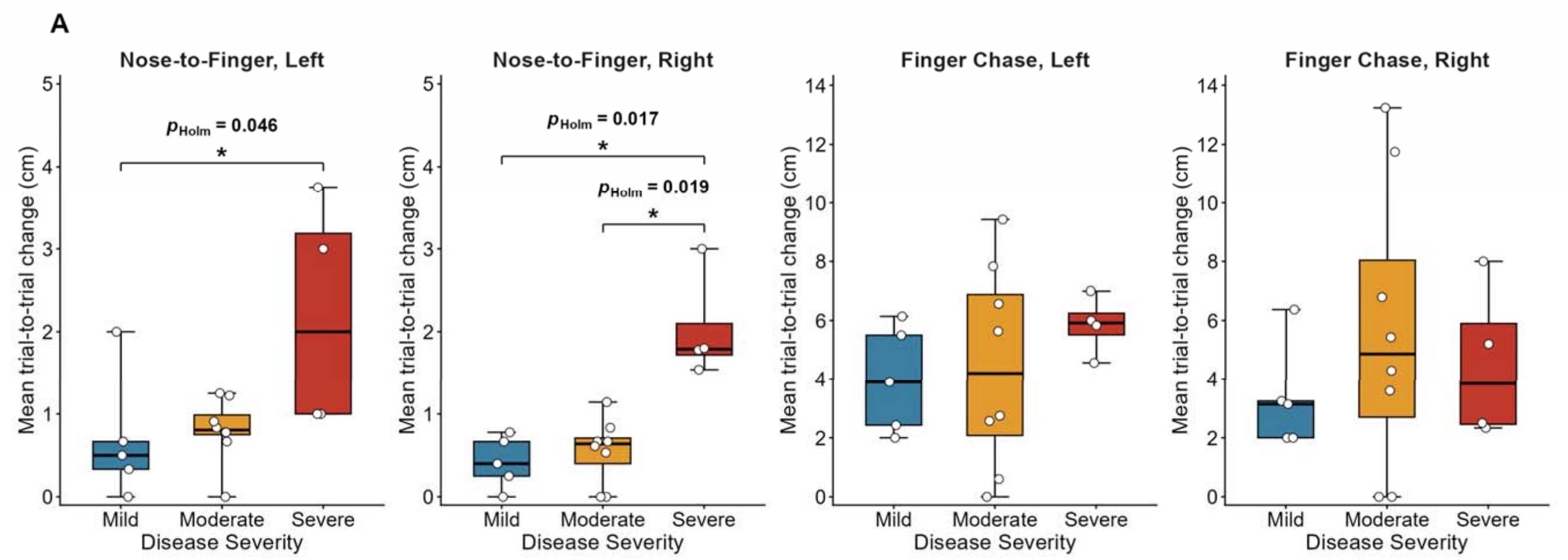
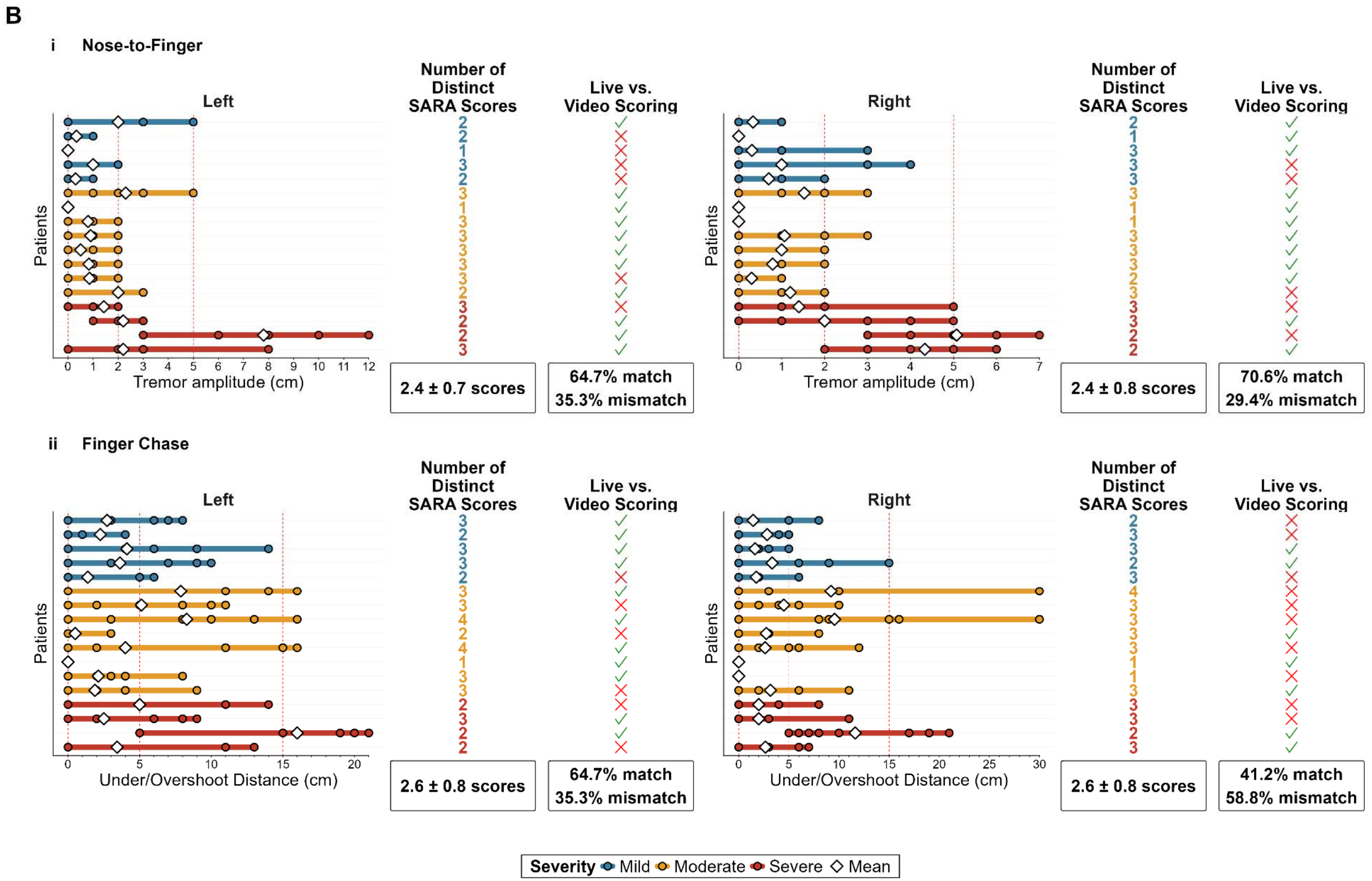
Trial-to-trial variability in SARA upper-limb items. **(A)** Mean trial-to-trial change (the mean absolute difference, in cm, between consecutive excursions) in SARA nose-to-finger (tremor amplitude) and finger chase (under/overshoot distance) for the left and right hands, by disease severity based on FARS functional stage (mild, moderate, severe). Boxes show the median and interquartile range, and points show individual patients. Group differences were tested with the Kruskal-Wallis test followed by Dunn’s post hoc comparisons with Holm correction; significant pairwise comparisons are marked with Holm-adjusted p values (* = *p*_Holm_ < .05). **(B)** Individual excursions for each patient in **(i)** SARA nose-to-finger and **(ii)** SARA finger chase, for the left and right hands, with one row per patient, colored by severity. Diamonds represent the mean across excursions. Circles show individual excursion values; when a patient had the same value on more than one excursion, a single circle is shown. Each patient’s row is colored by FARS severity group: mild (blue), moderate (yellow) and severe (red), with the same colors used for the number of distinct SARA column. Red dashed lines mark the SARA score cut-offs (nose-to-finger: 0, 2 and 5 cm; finger chase: 0, 5 and 15 cm). For each hand, the adjacent columns show the number of different SARA scores obtained across that patient’s excursions (mean ± SD below) and whether the live and video-derived SARA scores matched (⍰) or mismatched (⍰), with the overall percentage of matches and mismatches below.

Corresponding trial-to-trial changes in SARA scores were 0.59 ± 0.42 points (left) and 0.58 ± 0.40 points (right).

The mean number of distinct SARA scores across trials was 2.4 ± 0.7 for the left hand and 2.4 ± 0.8 for the right hand (Table 1A).

### SARA Finger Chase Trial-to-Trial Variability

For FC, mean undershoot/overshoot distance was 4.16 ± 3.79 cm (left) and 3.59 ± 3.34 cm (right) (Table 1B; Figure 1B). Patients deviated from this mean by 3.98 ± 1.90 cm (left) and 4.09 ± 3.10 cm (right), with under/overshoot distance ranges of 10.18 ± 4.98 cm (left) and 10.71 ± 8.46 cm (right) (Table 1B). Mean absolute trial-to-trial change in under/overshoot distance was 4.63 ± 2.60 cm (left) and 4.70 ± 3.68 cm (right), with corresponding mean trial-to-trial SARA score changes of 0.92 ± 0.52 (left) and 0.94 ± 0.50 (right) (Table 1B). Mean trial-to-trial under/overshoot distance change did not differ significantly across severity groups (left hand: 3.99 ± 1.82 cm mild, 4.42 ± 3.45 cm moderate, 5.85 ± 1.00 cm severe; right hand: 3.35 ± 1.79 cm mild, 5.64 ± 4.87 cm moderate, 4.51 ± 2.67 cm severe; Table 1D; Figure 1A; Supplementary Table S1B).

The mean number of distinct SARA scores across trials was 2.6 ± 0.8 for both left and right hands (Table 1B).

Individual plots of all excursions for NTF and FC for each patient are in Supplementary Figure S2. Individual patients’ values (mean, SD, range, and trial-to-trial change) are in Supplementary Tables S2.

### Discrepancies in Scoring Live vs Retrospective Video with Grid Overlay

In NTF, live and video scores matched in 64.7% of patients (left hand) and 70.6% (right). The live score was higher than the video score in 23.5% of patients (left) and 17.6% (right), and lower in 11.8% (both hands) (Supplementary Figure S3A).

For FC, SARA instructs the examiner to perform 5 consecutive fast-pointing movements and score them based on the last 3 trials. We compared match rates between scoring as instructed and scoring based on all trials. Match rates were 64.7% (left) and 41.2% (right) following the instructions (Supplementary Figure S3B and Supplementary Table S3) and 82.4% (left) and 70.6% (right) using the all trials method (Supplementary Table S3).

## Discussion

We scored SARA NTF and FC tasks in 17 patients with cerebellar ataxia live and by retrospective video review using a 1-cm calibrated grid overlay. We found marked trial-to-trial variability at all levels of ataxia severity. Most patients (NTF: 14/17, FC 15/17) had at least two distinct SARA scores across excursions. Trial-to-trial variability was higher for NTF in the severe group, whereas it was high across all severity stages for FC.

Live scores were discordant with grid-guided video measurements, ranging from ∼30% to ∼60% for the different tasks, and more pronounced for FC. Live scoring often overestimated impairment relative to video review, perhaps reflecting the high demand of judging and averaging centimeter-scale distances in real time.

The physiological basis of trial-to-trial variability has relevance for our findings. The cerebellar internal feedforward model holds that cerebellum predicts consequences of motor commands^7– 9^, and therefore reaching is impaired in patients with cerebellar ataxia because they are unable to adjust the predictive components of their movements. They fail to use error information from one reach to update the next^10,11^, producing variability of repeated movements characteristic of dysmetria^3^. This may explain why our patients received different scores from one trial to the next within the same examination. Scoring tremor amplitude (NTF), or under/overshoot distance (FC) on a centimeter ordinal scale based on three excursions thus poses the risk of an unrepresentative snapshot of a patient’s level of impairment, compounded by the suboptimal agreement between live and video-derived scores from the 1-cm grid. Granular anchor-based descriptors^12,13^ together with digital biomarkers that capture ecologically valid metrics of motor control^14–20^ may be necessary for the fine-grained quantitation of cerebellar ataxia necessary for clinical trial design.

### Limitations

We recognized potential for perspective distortion of measurement when video recording at 45 degrees, which we mitigated by the fixed location of the camera relative to the grid, and close approximation of the examiner and patient’s fingers to the grid. Differences reported in the severity subgroups were significant; however, the cohorts were small, and results therefore should be regarded as exploratory.

## Conclusions

Trial-to-trial variability and discrepancies between live and video scoring of NTF and FC pose a challenge to the reliability of centimeter-based ordinal scoring of upper-limb dysmetria in cross-sectional and longitudinal studies.

## Author Roles

(1) Research Project: A. Conception, B. Organization, C. Execution; (2) Statistical Analysis: A. Design, B. Execution, C. Review and Critique; (3) Manuscript Preparation: A. Writing of the First Draft, B. Review and Critique.

D.A.: 1B, 1C, 2A, 2B, 3A, 3B

J.D.S.: 1A, 1B, 1C, 2C, 3A, 3B

## Financial Disclosures for the previous 12⍰months

JDS is site PI for Biohaven Pharmaceuticals clinical trials NCT03701399 and NCT02960893 and IntraBio Inc clinical trials NCT03759678 and NCT07221292; consults for Biohaven Pharmaceuticals, IntraBio Inc, and Pretzel Therapeutics; receives royalties from Oxford University Press, Elsevier, MacKeith Press, and Springer; and is the inventor of the Brief Ataxia Rating Scale, Cerebellar Cognitive Affective/Schmahmann Syndrome Scale, the Patient Reported Outcome Measure of Ataxia, and the Cerebellar Neuropsychiatry Rating Scale which are licensed to the Schmahmann Lab and the Massachusetts General Hospital Corporation.

## Data Availability Statement

The data that support the findings of this study are available from the corresponding author upon reasonable request.

## Ethical Compliance Statement

This study was approved by the Mass General Brigham Institutional Review Board (IRB) on March 18, 2026 (IRB number 2026P000653). Informed consent was not required for this study. As part of the recruitment process, patients were provided with a fact sheet describing the study. Patients provided implied consent by choosing to participate or declining the invitation. We confirm that we have read the Journal’s position on issues involved in ethical publication and affirm that this work is consistent with those guidelines.

---

**Table 1A.** Trial-level variability in the NTF task.

| **NTF Trial-to-Trial Variability Metrics (N = 17)** | **Left Hand** | **Right Hand** |
|---|---|---|
| Age (years) | 50.8 (17.4) |  |
| FARS Functional Stage Score | 2.74 (1.31) |  |
| N Trials | 8.0 (2.2) | 12.2 (4.4) |
| DATA DERIVED FROM THE LIVE SCORING |  |  |
| Live SARA Score | 1.41 (0.87) | 1.12 (0.78) |
| DATA DERIVED FROM THE VIDEO REVIEW |  |  |
| Number of Distinct SARA Scores | 2.4 (0.7) | 2.4 (0.8) |
| Video SARA Score | 1.05 (0.74) | 0.88 (0.73) |
| Video SARA Score SD | 0.61 (0.30) | 0.54 (0.29) |
| Video SARA Score Range | 1.59 (0.80) | 1.41 (0.80) |
| Mean Tremor Amplitude (cm) | 1.50 (1.80) | 1.24 (1.43) |
| Tremor Amplitude SD (cm) | 0.99 (0.87) | 0.81 (0.57) |
| Tremor Amplitude Range (cm) | 2.82 (2.53) | 2.41 (1.66) |
| Trial-to-Trial Tremor Amplitude Change (cm) | 1.10 (0.99) | 0.86 (0.79) |
| Trial-to-Trial Tremor Amplitude Change SD (cm) | 0.78 (0.54) | 0.74 (0.48) |
| Trial-to-Trial SARA Score Change | 0.59 (0.42) | 0.58 (0.40) |
| Trial-to-Trial SARA Score Change SD | 0.56 (0.25) | 0.51 (0.28) |

Because the number of trials varied across patients and between hands, the mean and SD of the number of trials are reported for each hand. Age and FARS Functional Stage are patient-level variables that do not vary by hand and are therefore reported once across both columns.
N Trials: mean of the number of excursions performed by each hand.
Live SARA Score: mean final SARA score generated at the time of the live administration.
The remainder of the data in the table are derived from video-review.
Number of Distinct SARA Scores: how many different SARA scores (0 to 3) a patient’s excursions produced when each excursion was scored.
Video SARA Score: mean value of individual patient’s mean per-excursion SARA scores across all of a patient’s excursions when each excursion was scored.
Video SARA Score SD: mean of individual patients’ SDs of their per-excursion SARA scores.
Video SARA Score Range: mean of individual patient’s range for per-excursion SARA scores.
Mean Tremor Amplitude (cm): mean value of individual patients’ mean tremor amplitudes, with each patient’s mean calculated across all trials.
Tremor Amplitude SD (cm): mean of the individual patient SDs, with each patient’s SD calculated across trials.
Tremor Amplitude Range (cm): within-patient maximum minus minimum value, with the mean across patients reported.
Trial-to-Trial Tremor Amplitude Change (cm): mean of individual patients’ absolute difference in tremor amplitude between consecutive trials.
Trial-to-Trial Tremor Amplitude Change SD (cm): mean of individual patients’ SDs of the absolute differences between consecutive trials.
Trial-to-Trial SARA Score Change: mean of individual patients’ mean absolute difference in trial-level SARA scores between consecutive trials.
Trial-to-Trial SARA Score Change SD: mean of individual patients’ SDs of the absolute differences in trial-level SARA score between consecutive trials.

**Table 1B.** Trial-level variability in the FC task.

| FC Trial-to-Trial Variability Metrics (N = 17) | Left Hand | Right Hand |
|---|---|---|
| Age (years) | 50.8 (17.4) |  |
| FARS Functional Stage Score | 2.74 (1.31) |  |
| N Trials | 8.2 (1.6) | 7.8 (1.5) |
| DATA DERIVED FROM THE LIVE SCORING |  |  |
| Live SARA Score | 1.35 (0.70) | 0.88 (0.60) |
| DATA DERIVED FROM THE VIDEO REVIEW |  |  |
| Number of Distinct SARA Scores | 2.6 (0.8) | 2.6 (0.8) |
| Video SARA Score | 0.91 (0.63) | 0.82 (0.57) |
| Video SARA Score SD | 0.81 (0.31) | 0.78 (0.34) |
| Video SARA Score Range | 1.88 (0.78) | 1.82 (0.81) |
| Mean Under/Overshoot Distance (cm) | 4.16 (3.79) | 3.59 (3.34) |
| Under/Overshoot Distance SD (cm) | 3.98 (1.90) | 4.09 (3.10) |
| Under/Overshoot Distance Range (cm) | 10.18 (4.98) | 10.71 (8.46) |
| Trial-to-Trial Under/Overshoot Distance Change (cm) | 4.63 (2.60) | 4.70 (3.68) |
| Trial-to-Trial Under/Overshoot Distance Change SD (cm) | 3.58 (1.72) | 3.67 (2.75) |
| Trial-to-Trial SARA Score Change | 0.92 (0.52) | 0.94 (0.50) |
| Trial-to-Trial SARA Score Change SD | 0.77 (0.29) | 0.66 (0.30) |

Because the number of trials varied across patients and between hands, the mean and SD of the number of trials are reported for each hand. Age and FARS Functional Stage are patient-level variables that do not vary by hand and are therefore reported once across both columns.
N Trials: mean of the number of excursions performed by each hand.
Live SARA Score: mean final SARA score generated at the time of the live administration.
The remainder of the data in the table are derived from video-review.
Number of Distinct SARA Scores: how many different SARA scores (0 to 3) a patient’s excursions produced when each excursion was scored.
Video SARA Score: mean value of individual patient’s mean per-excursion SARA scores across all of a patient’s excursions when each excursion was scored.
Video SARA Score SD: mean of individual patients’ SDs of their per-excursion SARA scores.
Video SARA Score Range: mean of individual patient’s range for per-excursion SARA scores.
Mean Under/Overshoot Distance (cm): mean value of individual patients’ mean under/overshoot distances, with each patient’s mean calculated across all trials.
Under/Overshoot Distance SD (cm): mean of the individual patient SDs, with each patient’s SD calculated across trials.
Under/Overshoot Distance Range (cm): within-patient maximum minus minimum value, with the mean across patients reported.
Trial-to-Trial Under/Overshoot Distance Change (cm): mean of individual patients’ absolute difference in under/overshoot distance between consecutive trials.
Trial-to-Trial Under/Overshoot Distance Change SD (cm): mean of individual patients’ SDs of the absolute differences between consecutive trials.
Trial-to-Trial SARA Score Change: mean of individual patients’ mean absolute difference in trial-level SARA scores between consecutive trials.
Trial-to-Trial SARA Score Change SD: mean of individual patients’ SDs of the absolute differences in trial-level SARA score between consecutive trials.

**Table 1C.**
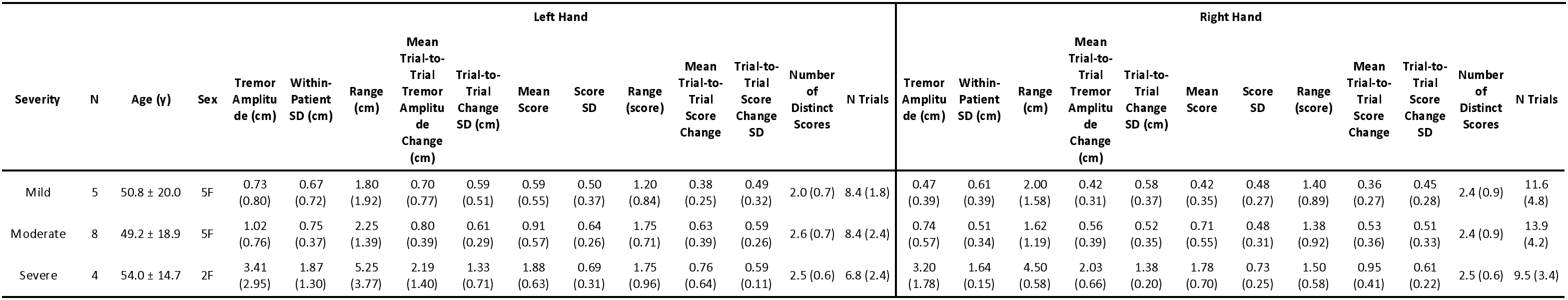
Trial-level variability of NTF by severity group.

| Severity | N | Age (y) | Sex | Left Hand |  |  |  |  |  |  |  |  |  |  |  | Right Hand |  |  |  |  |  |  |  |  |  |  |  |
|---|---|---|---|---|---|---|---|---|---|---|---|---|---|---|---|---|---|---|---|---|---|---|---|---|---|---|---|
|  |  |  |  | Tremor Amplitude (cm) | Within-Patient SD (cm) | Range (cm) | Mean Trial-to-Trial Tremor Amplitude Change (cm) | Trial-to-Trial Change SD (cm) | Mean Score | Score SD | Range (score) | Mean Trial-to-Trial Score Change | Trial-to-Trial Score Change SD | Number of Distinct Scores | N Trials | Tremor Amplitude (cm) | Within-Patient SD (cm) | Range (cm) | Mean Trial-to-Trial Tremor Amplitude Change (cm) | Trial-to-Trial Change SD (cm) | Mean Score | Score SD | Range (score) | Mean Trial-to-Trial Score Change | Trial-to-Trial Score Change SD | Number of Distinct Scores | N Trials |
| Mild | 5 | 50.8 ± 20.0 | 5F | 0.73 (0.80) | 0.67 (0.72) | 1.80 (1.92) | 0.70 (0.77) | 0.59 (0.51) | 0.59 (0.55) | 0.50 (0.37) | 1.20 (0.84) | 0.38 (0.25) | 0.49 (0.32) | 2.0 (0.7) | 8.4 (1.8) | 0.47 (0.39) | 0.61 (0.39) | 2.00 (1.58) | 0.42 (0.31) | 0.58 (0.37) | 0.42 (0.35) | 0.48 (0.27) | 1.40 (0.89) | 0.36 (0.27) | 0.45 (0.28) | 2.4 (0.9) | 11.6 (4.8) |
| Moderate | 8 | 49.2 ± 18.9 | 5F | 1.02 (0.76) | 0.75 (0.37) | 2.25 (1.39) | 0.80 (0.39) | 0.61 (0.29) | 0.91 (0.57) | 0.64 (0.26) | 1.75 (0.71) | 0.63 (0.39) | 0.59 (0.26) | 2.6 (0.7) | 8.4 (2.4) | 0.74 (0.57) | 0.51 (0.34) | 1.62 (1.19) | 0.56 (0.39) | 0.52 (0.35) | 0.71 (0.55) | 0.48 (0.31) | 1.38 (0.92) | 0.53 (0.36) | 0.51 (0.33) | 2.4 (0.9) | 13.9 (4.2) |
| Severe | 4 | 54.0 ± 14.7 | 2F | 3.41 (2.95) | 1.87 (1.30) | 5.25 (3.77) | 2.19 (1.40) | 1.33 (0.71) | 1.88 (0.63) | 0.69 (0.31) | 1.75 (0.96) | 0.76 (0.64) | 0.59 (0.11) | 2.5 (0.6) | 6.8 (2.4) | 3.20 (1.78) | 1.64 (0.15) | 4.50 (0.58) | 2.03 (0.66) | 1.38 (0.20) | 1.78 (0.70) | 0.73 (0.25) | 1.50 (0.58) | 0.95 (0.41) | 0.61 (0.22) | 2.5 (0.6) | 9.5 (3.4) |

**Table 1D.**
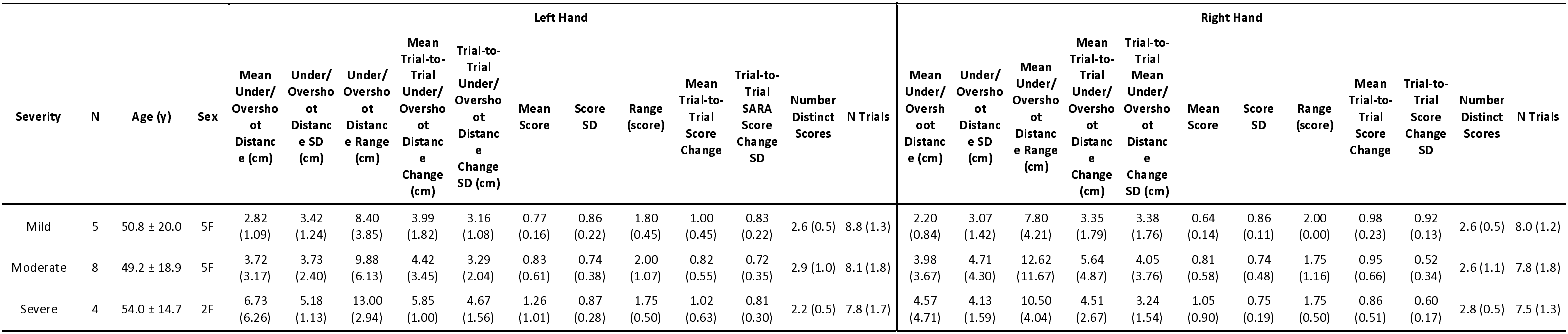
Trial-level variability of FC task by severity group.

|  |  |  |  | Left Hand |  |  |  |  |  |  |  |  |  |  |  | Right Hand |  |  |  |  |  |  |  |  |  |  |  |
|---|---|---|---|---|---|---|---|---|---|---|---|---|---|---|---|---|---|---|---|---|---|---|---|---|---|---|---|
| Severity | N | Age (y) | Sex | Mean Under/Overshoot Distance (cm) | Under/Overshoot Distance SD (cm) | Under/Overshoot Distance Range (cm) | Mean Trial-to-Trial Under/Overshoot Distance Change (cm) | Trial-to-Trial Under/Overshoot Distance Change SD (cm) | Mean Score | Score SD | Range (score) | Mean Trial-to-Trial Score Change | Trial-to-Trial SARA Score Change SD | Number Distinct Scores | N Trials | Mean Under/Overshoot Distance (cm) | Under/Overshoot Distance SD (cm) | Mean Under/Overshoot Distance Range (cm) | Mean Trial-to-Trial Under/Overshoot Distance Change (cm) | Trial-to-Trial Mean Under/Overshoot Distance Change SD (cm) | Mean Score | Score SD | Range (score) | Mean Trial-to-Trial Score Change | Trial-to-Trial Score Change SD | Number Distinct Scores | N Trials |
| Mild | 5 | 50.8 ± 20.0 | 5F | 2.82 (1.09) | 3.42 (1.24) | 8.40 (3.85) | 3.99 (1.82) | 3.16 (1.08) | 0.77 (0.16) | 0.86 (0.22) | 1.80 (0.45) | 1.00 (0.45) | 0.83 (0.22) | 2.6 (0.5) | 8.8 (1.3) | 2.20 (0.84) | 3.07 (1.42) | 7.80 (4.21) | 3.35 (1.79) | 3.38 (1.76) | 0.64 (0.14) | 0.86 (0.11) | 2.00 (0.00) | 0.98 (0.23) | 0.92 (0.13) | 2.6 (0.5) | 8.0 (1.2) |
| Moderate | 8 | 49.2 ± 18.9 | 5F | 3.72 (3.17) | 3.73 (2.40) | 9.88 (6.13) | 4.42 (3.45) | 3.29 (2.04) | 0.83 (0.61) | 0.74 (0.38) | 2.00 (1.07) | 0.82 (0.55) | 0.72 (0.35) | 2.9 (1.0) | 8.1 (1.8) | 3.98 (3.67) | 4.71 (4.30) | 12.62 (11.67) | 5.64 (4.87) | 4.05 (3.76) | 0.81 (0.58) | 0.74 (0.48) | 1.75 (1.16) | 0.95 (0.66) | 0.52 (0.34) | 2.6 (1.1) | 7.8 (1.8) |
| Severe | 4 | 54.0 ± 14.7 | 2F | 6.73 (6.26) | 5.18 (1.13) | 13.00 (2.94) | 5.85 (1.00) | 4.67 (1.56) | 1.26 (1.01) | 0.87 (0.28) | 1.75 (0.50) | 1.02 (0.63) | 0.81 (0.30) | 2.2 (0.5) | 7.8 (1.7) | 4.57 (4.71) | 4.13 (1.59) | 10.50 (4.04) | 4.51 (2.67) | 3.24 (1.54) | 1.05 (0.90) | 0.75 (0.19) | 1.75 (0.50) | 0.86 (0.51) | 0.60 (0.17) | 2.8 (0.5) | 7.5 (1.3) |

