## Supplementary Tables for "Challenges in Quantifying Upper Extremity Dysmetria in the Scale for the Assessment and Rating of Ataxia (SARA)"

**Supplementary Table S1A. Nose-to-Finger (NTF).** Kruskal-Wallis rank-sum tests (df = 2) and Dunn’s post hoc comparisons of the NTF measures across FARS-Func severity groups (Mild (n = 5), Moderate (n = 8) and Severe (n = 4)). Post hoc *p*-values are Holm-adjusted, computed separately for each hand. *p* = unadjusted *p*-value, *p*_Holm_ = Holm-adjusted *p*-value, adjusted across the 3 pairwise comparisons. Bold = *p*_Holm_ < .05.

| **Hand** | **Measure** | **Kruskal-Wallis H (df = 2)** | **Kruskal-Wallis *p*** | **Mild vs Moderate** | **Mild vs Severe** | **Moderate vs Severe** |
| --- | --- | --- | --- | --- | --- | --- |
|  | Age (years) | 0.15 | 0.928 | *p* = 0.852, *p*_Holm_ = 1.000 | *p* = 0.848, *p*_Holm_ = 1.000 | *p* = 0.701, *p*_Holm_ = 1.000 |
| Left | Live SARA Score | 5.18 | 0.075 | *p* = 0.727, *p*_Holm_ = 0.727 | *p* = 0.036, *p*_Holm_ = 0.107 | *p* = 0.048, *p*_Holm_ = 0.107 |
|  | N Trials | 1.65 | 0.438 | *p* = 0.979, *p*_Holm_ = 0.979 | *p* = 0.279, *p*_Holm_ = 0.679 | *p* = 0.226, *p*_Holm_ = 0.679 |
|  | Mean Tremor Amplitude (cm) | 6.12 | 0.047 | *p* = 0.503, *p*_Holm_ = 0.503 | *p* = 0.017, *p*_Holm_ = 0.052 | *p* = 0.047, *p*_Holm_ = 0.094 |
|  | Tremor Amplitude SD (cm) | 5.52 | 0.063 | *p* = 0.289, *p*_Holm_ = 0.289 | *p* = 0.019, *p*_Holm_ = 0.057 | *p* = 0.114, *p*_Holm_ = 0.229 |
|  | Tremor Amplitude Range (cm) | 3.75 | 0.153 | *p* = 0.332, *p*_Holm_ = 0.446 | *p* = 0.053, *p*_Holm_ = 0.158 | *p* = 0.223, *p*_Holm_ = 0.446 |
|  | Trial-to-Trial Tremor Amplitude Change (cm) | 6.04 | 0.049 | *p* = 0.347, *p*_Holm_ = 0.347 | ***p* = 0.015, *p*_Holm_ = 0.046** | *p* = 0.075, *p*_Holm_ = 0.149 |
|  | Trial-to-Trial Tremor Amplitude Change SD (cm) | 4.74 | 0.094 | *p* = 0.378, *p*_Holm_ = 0.378 | *p* = 0.031, *p*_Holm_ = 0.093 | *p* = 0.123, *p*_Holm_ = 0.246 |
|  | Trial-to-Trial SARA Score Change | 1.12 | 0.570 | *p* = 0.316, *p*_Holm_ = 0.947 | *p* = 0.415, *p*_Holm_ = 0.947 | *p* = 0.968, *p*_Holm_ = 0.968 |
|  | Trial-to-Trial SARA Score Change SD | 1.18 | 0.554 | *p* = 0.288, *p*_Holm_ = 0.865 | *p* = 0.719, *p*_Holm_ = 1.000 | *p* = 0.552, *p*_Holm_ = 1.000 |
|  | SARA Score SD | 1.03 | 0.599 | *p* = 0.324, *p*_Holm_ = 0.971 | *p* = 0.478, *p*_Holm_ = 0.971 | *p* = 0.887, *p*_Holm_ = 0.971 |
|  | SARA Score Range | 1.98 | 0.372 | *p* = 0.164, *p*_Holm_ = 0.491 | *p* = 0.373, *p*_Holm_ = 0.745 | *p* = 0.749, *p*_Holm_ = 0.749 |
|  | Number of Distinct SARA Scores | 2.94 | 0.230 | *p* = 0.087, *p*_Holm_ = 0.262 | *p* = 0.317, *p*_Holm_ = 0.634 | *p* = 0.621, *p*_Holm_ = 0.634 |
| Right | Live SARA Score | 7.10 | 0.029 | *p* = 0.900, *p*_Holm_ = 0.900 | ***p* = 0.020, *p*_Holm_ = 0.044** | ***p* = 0.015, *p*_Holm_ = 0.044** |
|  | N Trials | 2.98 | 0.225 | *p* = 0.340, *p*_Holm_ = 0.680 | *p* = 0.467, *p*_Holm_ = 0.680 | *p* = 0.092, *p*_Holm_ = 0.276 |
|  | Mean Tremor Amplitude (cm) | 8.51 | 0.014 | *p* = 0.505, *p*_Holm_ = 0.505 | ***p* = 0.006, *p*_Holm_ = 0.017** | ***p* = 0.016, *p*_Holm_ = 0.032** |
|  | Tremor Amplitude SD (cm) | 9.04 | 0.011 | *p* = 0.571, *p*_Holm_ = 0.571 | *p* = 0.026, *p*_Holm_ = 0.053 | ***p* = 0.003, *p*_Holm_ = 0.009** |
|  | Tremor Amplitude Range (cm) | 8.43 | 0.015 | *p* = 0.679, *p*_Holm_ = 0.679 | *p* = 0.026, *p*_Holm_ = 0.051 | ***p* = 0.005, *p*_Holm_ = 0.014** |
|  | Trial-to-Trial Tremor Amplitude Change (cm) | 8.96 | 0.011 | *p* = 0.650, *p*_Holm_ = 0.650 | ***p* = 0.006, *p*_Holm_ = 0.017** | ***p* = 0.009, *p*_Holm_ = 0.019** |
|  | Trial-to-Trial Tremor Amplitude Change SD (cm) | 8.75 | 0.013 | *p* = 0.865, *p*_Holm_ = 0.865 | ***p* = 0.015, *p*_Holm_ = 0.030** | ***p* = 0.005, *p*_Holm_ = 0.014** |
|  | Trial-to-Trial SARA Score Change | 4.66 | 0.097 | *p* = 0.337, *p*_Holm_ = 0.337 | *p* = 0.031, *p*_Holm_ = 0.094 | *p* = 0.144, *p*_Holm_ = 0.287 |
|  | Trial-to-Trial SARA Score Change SD | 0.36 | 0.836 | *p* = 0.623, *p*_Holm_ = 1.000 | *p* = 0.584, *p*_Holm_ = 1.000 | *p* = 0.887, *p*_Holm_ = 1.000 |
|  | SARA Score SD | 1.07 | 0.587 | *p* = 0.714, *p*_Holm_ = 0.931 | *p* = 0.310, *p*_Holm_ = 0.931 | *p* = 0.441, *p*_Holm_ = 0.931 |
|  | SARA Score Range | 0.00 | 1.000 | *p* = 1.000, *p*_Holm_ = 1.000 | *p* = 1.000, *p*_Holm_ = 1.000 | *p* = 1.000, *p*_Holm_ = 1.000 |
|  | Number of Distinct SARA Scores | 0.00 | 1.000 | *p* = 1.000, *p*_Holm_ = 1.000 | *p* = 1.000, *p*_Holm_ = 1.000 | *p* = 1.000, *p*_Holm_ = 1.000 |

**Supplementary Table S1B. Finger Chase (FC).** Kruskal-Wallis tests and Dunn’s post hoc comparisons of the FC measures across FARS-Func severity groups (Mild (n = 5), Moderate (n = 8) and Severe (n = 4)). Post hoc *p*-values are Holm-adjusted, computed separately for each hand. *p* = unadjusted *p*-value, *p*_Holm_ = Holm-adjusted *p*-value, adjusted across the 3 pairwise comparisons. Bold = *p*_Holm_ < .05.

| **Hand** | **Measure** | **Kruskal-Wallis H (df = 2)** | **Kruskal-Wallis *p*** | **Mild vs Moderate** | **Mild vs Severe** | **Moderate vs Severe** |
| --- | --- | --- | --- | --- | --- | --- |
|  | Age (years) | 0.15 | 0.928 | *p* = 0.852, *p*_Holm_ = 1.000 | *p* = 0.848, *p*_Holm_ = 1.000 | *p* = 0.701, *p*_Holm_ = 1.000 |
| Left | Live SARA Score | 1.12 | 0.571 | *p* = 0.805, *p*_Holm_ = 0.942 | *p* = 0.314, *p*_Holm_ = 0.942 | *p* = 0.382, *p*_Holm_ = 0.942 |
|  | N Trials | 1.70 | 0.427 | *p* = 0.291, *p*_Holm_ = 0.684 | *p* = 0.228, *p*_Holm_ = 0.684 | *p* = 0.737, *p*_Holm_ = 0.737 |
|  | Mean Under/Overshoot Distance (cm) | 1.34 | 0.511 | *p* = 0.808, *p*_Holm_ = 0.824 | *p* = 0.275, *p*_Holm_ = 0.824 | *p* = 0.332, *p*_Holm_ = 0.824 |
|  | Under/Overshoot Distance SD (cm) | 2.78 | 0.249 | *p* = 0.602, *p*_Holm_ = 0.602 | *p* = 0.104, *p*_Holm_ = 0.313 | *p* = 0.196, *p*_Holm_ = 0.392 |
|  | Under/Overshoot Distance Range (cm) | 2.06 | 0.356 | *p* = 0.428, *p*_Holm_ = 0.807 | *p* = 0.151, *p*_Holm_ = 0.453 | *p* = 0.404, *p*_Holm_ = 0.807 |
|  | Trial-to-Trial Under/Overshoot Distance Change (cm) | 1.62 | 0.445 | *p* = 0.561, *p*_Holm_ = 0.792 | *p* = 0.204, *p*_Holm_ = 0.613 | *p* = 0.396, *p*_Holm_ = 0.792 |
|  | Trial-to-Trial Under/Overshoot Distance Change SD (cm) | 2.17 | 0.338 | *p* = 1.000, *p*_Holm_ = 1.000 | *p* = 0.210, *p*_Holm_ = 0.508 | *p* = 0.169, *p*_Holm_ = 0.508 |
|  | Trial-to-Trial SARA Score Change | 0.77 | 0.681 | *p* = 0.504, *p*_Holm_ = 1.000 | *p* = 0.894, *p*_Holm_ = 1.000 | *p* = 0.442, *p*_Holm_ = 1.000 |
|  | Trial-to-Trial SARA Score Change SD | 0.41 | 0.816 | *p* = 0.552, *p*_Holm_ = 1.000 | *p* = 0.906, *p*_Holm_ = 1.000 | *p* = 0.671, *p*_Holm_ = 1.000 |
|  | SARA Score SD | 0.66 | 0.719 | *p* = 0.602, *p*_Holm_ = 1.000 | *p* = 0.796, *p*_Holm_ = 1.000 | *p* = 0.442, *p*_Holm_ = 1.000 |
|  | SARA Score Range | 0.86 | 0.650 | *p* = 0.466, *p*_Holm_ = 1.000 | *p* = 0.914, *p*_Holm_ = 1.000 | *p* = 0.426, *p*_Holm_ = 1.000 |
|  | Number of Distinct SARA Scores | 2.31 | 0.315 | *p* = 0.484, *p*_Holm_ = 0.867 | *p* = 0.434, *p*_Holm_ = 0.867 | *p* = 0.131, *p*_Holm_ = 0.393 |
| Right | Live SARA Score | 1.55 | 0.460 | *p* = 0.238, *p*_Holm_ = 0.715 | *p* = 0.338, *p*_Holm_ = 0.715 | *p* = 0.962, *p*_Holm_ = 0.962 |
|  | N Trials | 0.46 | 0.794 | *p* = 0.588, *p*_Holm_ = 1.000 | *p* = 0.527, *p*_Holm_ = 1.000 | *p* = 0.851, *p*_Holm_ = 1.000 |
|  | Mean Under/Overshoot Distance (cm) | 0.90 | 0.637 | *p* = 0.375, *p*_Holm_ = 1.000 | *p* = 0.451, *p*_Holm_ = 1.000 | *p* = 1.000, *p*_Holm_ = 1.000 |
|  | Under/Overshoot Distance SD (cm) | 1.62 | 0.446 | *p* = 0.255, *p*_Holm_ = 0.765 | *p* = 0.281, *p*_Holm_ = 0.765 | *p* = 0.903, *p*_Holm_ = 0.903 |
|  | Under/Overshoot Distance Range (cm) | 1.37 | 0.505 | *p* = 0.293, *p*_Holm_ = 0.879 | *p* = 0.324, *p*_Holm_ = 0.879 | *p* = 0.919, *p*_Holm_ = 0.919 |
|  | Trial-to-Trial Under/Overshoot Distance Change (cm) | 1.19 | 0.550 | *p* = 0.277, *p*_Holm_ = 0.831 | *p* = 0.506, *p*_Holm_ = 1.000 | *p* = 0.777, *p*_Holm_ = 1.000 |
|  | Trial-to-Trial Under/Overshoot Distance Change SD (cm) | 0.04 | 0.982 | *p* = 0.876, *p*_Holm_ = 1.000 | *p* = 0.988, *p*_Holm_ = 1.000 | *p* = 0.871, *p*_Holm_ = 1.000 |
|  | Trial-to-Trial SARA Score Change | 0.75 | 0.688 | *p* = 0.876, *p*_Holm_ = 1.000 | *p* = 0.520, *p*_Holm_ = 1.000 | *p* = 0.395, *p*_Holm_ = 1.000 |
|  | Trial-to-Trial SARA Score Change SD | 7.35 | **0.025** | ***p* = 0.016, *p*_Holm_ = 0.048** | ***p* = 0.020, *p*_Holm_ = 0.048** | *p* = 0.761, *p*_Holm_ = 0.761 |
|  | SARA Score SD | 0.93 | 0.628 | *p* = 0.976, *p*_Holm_ = 1.000 | *p* = 0.421, *p*_Holm_ = 1.000 | *p* = 0.362, *p*_Holm_ = 1.000 |
|  | SARA Score Range | 0.41 | 0.813 | *p* = 0.914, *p*_Holm_ = 1.000 | *p* = 0.551, *p*_Holm_ = 1.000 | *p* = 0.580, *p*_Holm_ = 1.000 |
|  | Number of Distinct SARA Scores | 0.25 | 0.884 | *p* = 0.632, *p*_Holm_ = 1.000 | *p* = 0.716, *p*_Holm_ = 1.000 | *p* = 0.962, *p*_Holm_ = 1.000 |

**Supplementary Table S2A.** Individual patient data for the NTF task.

|  | | | | **Left Hand** | | | | | | | | **Right Hand** | | | | | | | |
| --- | --- | --- | --- | --- | --- | --- | --- | --- | --- | --- | --- | --- | --- | --- | --- | --- | --- | --- | --- |
| **ID** | **Age range (y)** | **Sex** | **Diagnosis** | **Tremor Amplitude Mean (SD) (cm)** | **Tremor Amplitude Range (cm)** | **Mean Trial-to-Trial Tremor Amplitude Change (cm)** | **Mean (SD) (score)** | **Range (score)** | **Mean Trial-to-Trial Score Change** | **Number of Distinct Scores** | **N Trials** | **Tremor Amplitude Mean (SD) (cm)** | **Tremor Amplitude Range (cm)** | **Mean Trial-to-Trial Tremor Amplitude Change (cm)** | **Mean (SD) (score)** | **Range (score)** | **Mean Trial-to-Trial Score Change** | **Number of Distinct Scores** | **N Trials** |
| 1 | 26–30 | F | SCA2 | 2.00 (1.90) | 5.00 | 2.00 | 1.33 (1.03) | 2 | 0.40 | 2 | 6 | 0.33 (0.52) | 1.00 | 0.40 | 0.33 (0.52) | 1 | 0.40 | 2 | 6 |
| 2 | 66–70 | M | SCA3 | 2.20 (0.84) | 2.00 | 1.00 | 1.80 (0.45) | 1 | 0.25 | 2 | 5 | 2.00 (1.85) | 5.00 | 3.00 | 1.25 (0.89) | 2 | 1.43 | 3 | 8 |
| 3 | 36–40 | F | SCA3 | 0.33 (0.50) | 1.00 | 0.50 | 0.33 (0.50) | 1 | 0.50 | 2 | 9 | 0.00 (0.00) | 0.00 | 0.00 | 0.00 (0.00) | 0 | 0.00 | 1 | 10 |
| 4 | 36–40 | M | SCA1 | 2.30 (1.34) | 5.00 | 1.22 | 1.70 (0.67) | 2 | 0.33 | 3 | 10 | 1.53 (0.84) | 3.00 | 0.83 | 1.42 (0.69) | 2 | 0.78 | 3 | 19 |
| 5 | 76–80 | F | SCA6 | 2.00 (1.00) | 3.00 | 0.83 | 1.71 (0.76) | 2 | 0.33 | 2 | 7 | 1.20 (0.63) | 2.00 | 0.67 | 1.20 (0.63) | 2 | 0.67 | 3 | 10 |
| 6 | 66–70 | F | SCA27B | 0.00 (0.00) | 0.00 | 0.00 | 0.00 (0.00) | 0 | 0.00 | 1 | 7 | 0.31 (0.85) | 3.00 | 0.25 | 0.23 (0.60) | 2 | 0.17 | 3 | 13 |
| 7 | 71–75 | F | SCA27B | 1.00 (0.47) | 2.00 | 0.33 | 1.00 (0.47) | 2 | 0.33 | 3 | 10 | 1.00 (1.00) | 4.00 | 0.78 | 0.84 (0.60) | 2 | 0.56 | 3 | 19 |
| 8 | 36–40 | F | SCA3 | 0.50 (0.71) | 2.00 | 0.78 | 0.50 (0.71) | 2 | 0.78 | 3 | 10 | 1.00 (0.63) | 2.00 | 0.53 | 1.00 (0.63) | 2 | 0.53 | 3 | 16 |
| 9 | 41–45 | F | SCA3 | 0.30 (0.48) | 1.00 | 0.67 | 0.30 (0.48) | 1 | 0.67 | 2 | 10 | 0.70 (0.67) | 2.00 | 0.67 | 0.70 (0.67) | 2 | 0.67 | 3 | 10 |
| 10 | 26–30 | M | SCA3 | 0.83 (0.72) | 2.00 | 0.91 | 0.83 (0.72) | 2 | 0.91 | 3 | 12 | 0.79 (0.63) | 2.00 | 0.61 | 0.79 (0.63) | 2 | 0.61 | 3 | 19 |
| 11 | 51–55 | M | SCA8 | 7.80 (3.49) | 9.00 | 3.75 | 2.80 (0.45) | 1 | 0.25 | 2 | 5 | 5.07 (1.49) | 4.00 | 1.54 | 2.43 (0.51) | 1 | 0.46 | 2 | 14 |
| 12 | 61–65 | F | SCA1 | 1.43 (0.79) | 2.00 | 1.00 | 1.43 (0.79) | 2 | 1.00 | 3 | 7 | 1.40 (1.58) | 5.00 | 1.78 | 1.10 (0.99) | 2 | 1.11 | 3 | 10 |
| 13 | 71–75 | F | SCA6 | 0.00 (0.00) | 0.00 | 0.00 | 0.00 (0.00) | 0 | 0.00 | 1 | 6 | 0.00 (0.00) | 0.00 | 0.00 | 0.00 (0.00) | 0 | 0.00 | 1 | 14 |
| 14 | 31–35 | F | Autoimmune-mediated cerebellar syndrome | 0.80 (0.84) | 2.00 | 1.25 | 0.80 (0.84) | 2 | 1.25 | 3 | 5 | 0.00 (0.00) | 0.00 | 0.00 | 0.00 (0.00) | 0 | 0.00 | 1 | 8 |
| 15 | 56–60 | F | SCA7 | 0.86 (0.69) | 2.00 | 0.67 | 0.86 (0.69) | 2 | 0.67 | 3 | 7 | 0.30 (0.48) | 1.00 | 0.67 | 0.30 (0.48) | 1 | 0.67 | 2 | 10 |
| 16 | 31–35 | F | SCA2 | 2.20 (2.35) | 8.00 | 3.00 | 1.50 (1.08) | 3 | 1.56 | 3 | 10 | 4.33 (1.63) | 4.00 | 1.80 | 2.33 (0.52) | 1 | 0.80 | 2 | 6 |
| 17 | 46–50 | M | SCA2 | 0.90 (0.74) | 2.00 | 0.78 | 0.90 (0.74) | 2 | 0.78 | 3 | 10 | 1.07 (0.88) | 3.00 | 1.14 | 1.00 (0.76) | 2 | 1.00 | 3 | 15 |

**Supplementary Table S2B.** Individual patient data for the FC task.

|  | | | | **Left Hand** | | | | | | | | **Right Hand** | | | | | | | |
| --- | --- | --- | --- | --- | --- | --- | --- | --- | --- | --- | --- | --- | --- | --- | --- | --- | --- | --- | --- |
| **ID** | **Age range (y)** | **Sex** | **Diagnosis** | **Under/Overshoot Distance Mean (SD) (cm)** | **Under/Overshoot Distance Range (cm)** | **Mean Trial-to-Trial Under/Overshoot Distance Change (cm)** | **Mean (SD) (score)** | **Range (score)** | **Mean Trial-to-Trial Score Change** | **Number of Distinct Scores** | **N Trials** | **Under/Overshoot Distance Mean (SD) (cm)** | **Under/Overshoot Distance Range (cm)** | **Mean Trial-to-Trial Under/Overshoot Distance Change (cm)** | **Mean (SD) (score)** | **Range (score)** | **Mean Trial-to-Trial Score Change** | **Number of Distinct Scores** | **N Trials** |
| 1 | 26–30 | F | SCA2 | 2.73 (3.35) | 8.00 | 3.90 | 0.82 (0.98) | 2 | 1.20 | 3 | 11 | 1.44 (2.96) | 8.00 | 3.25 | 0.44 (0.88) | 2 | 1.00 | 2 | 9 |
| 2 | 66–70 | M | SCA3 | 2.50 (3.69) | 9.00 | 4.56 | 0.70 (0.95) | 2 | 1.33 | 3 | 10 | 2.00 (4.12) | 11.00 | 2.33 | 0.43 (0.79) | 2 | 0.50 | 3 | 7 |
| 3 | 36–40 | F | SCA3 | 2.25 (1.91) | 4.00 | 2.00 | 0.75 (0.46) | 1 | 0.29 | 2 | 8 | 2.83 (2.23) | 5.00 | 2.00 | 0.83 (0.75) | 2 | 0.80 | 3 | 6 |
| 4 | 36–40 | M | SCA1 | 7.88 (6.75) | 16.00 | 9.43 | 1.38 (1.19) | 3 | 1.57 | 3 | 8 | 9.20 (12.19) | 30.00 | 11.75 | 1.40 (1.14) | 3 | 1.50 | 4 | 5 |
| 5 | 76–80 | F | SCA6 | 1.88 (3.23) | 9.00 | 2.57 | 0.50 (0.76) | 2 | 0.57 | 3 | 8 | 3.17 (4.49) | 11.00 | 6.80 | 0.83 (0.98) | 2 | 1.60 | 3 | 6 |
| 6 | 66–70 | F | SCA27B | 4.11 (4.91) | 14.00 | 5.50 | 0.89 (0.93) | 2 | 1.25 | 3 | 9 | 1.62 (1.92) | 5.00 | 2.00 | 0.62 (0.74) | 2 | 0.71 | 3 | 8 |
| 7 | 71–75 | F | SCA27B | 3.62 (4.37) | 10.00 | 6.14 | 0.88 (0.99) | 2 | 1.43 | 3 | 8 | 3.33 (5.50) | 15.00 | 6.38 | 0.67 (1.00) | 2 | 1.25 | 2 | 9 |
| 8 | 36–40 | F | SCA3 | 4.00 (6.19) | 16.00 | 5.64 | 0.83 (1.03) | 3 | 1.27 | 4 | 12 | 2.64 (3.75) | 12.00 | 3.60 | 0.82 (0.87) | 2 | 0.90 | 3 | 11 |
| 9 | 41–45 | F | SCA3 | 1.38 (2.56) | 6.00 | 2.43 | 0.50 (0.93) | 2 | 0.86 | 2 | 8 | 1.75 (2.71) | 6.00 | 3.14 | 0.62 (0.92) | 2 | 1.14 | 3 | 8 |
| 10 | 26–30 | M | SCA3 | 0.00 (0.00) | 0.00 | 0.00 | 0.00 (0.00) | 0 | 0.00 | 1 | 7 | 0.00 (0.00) | 0.00 | 0.00 | 0.00 (0.00) | 0 | 0.00 | 1 | 7 |
| 11 | 51–55 | M | SCA8 | 16.00 (4.96) | 16.00 | 6.00 | 2.75 (0.46) | 1 | 0.14 | 2 | 8 | 11.62 (6.37) | 16.00 | 8.00 | 2.38 (0.52) | 1 | 0.57 | 2 | 8 |
| 12 | 61–65 | F | SCA1 | 5.00 (6.20) | 14.00 | 7.00 | 1.00 (1.10) | 2 | 1.60 | 2 | 6 | 2.00 (2.83) | 8.00 | 2.50 | 0.56 (0.73) | 2 | 0.75 | 3 | 9 |
| 13 | 71–75 | F | SCA6 | 5.12 (4.29) | 11.00 | 6.57 | 1.38 (0.92) | 2 | 1.14 | 3 | 8 | 4.50 (3.96) | 10.00 | 5.43 | 1.12 (0.83) | 2 | 0.86 | 3 | 8 |
| 14 | 31–35 | F | Autoimmune-mediated cerebellar syndrome | 8.29 (5.50) | 16.00 | 7.83 | 1.71 (0.95) | 3 | 1.17 | 4 | 7 | 9.56 (9.80) | 30.00 | 13.25 | 1.56 (1.24) | 3 | 1.62 | 3 | 9 |
| 15 | 56–60 | F | SCA7 | 2.11 (2.67) | 8.00 | 2.75 | 0.67 (0.71) | 2 | 0.62 | 3 | 9 | 0.00 (0.00) | 0.00 | 0.00 | 0.00 (0.00) | 0 | 0.00 | 1 | 8 |
| 16 | 31–35 | F | SCA2 | 3.43 (5.88) | 13.00 | 5.83 | 0.57 (0.98) | 2 | 1.00 | 2 | 7 | 2.67 (3.20) | 7.00 | 5.20 | 0.83 (0.98) | 2 | 1.60 | 3 | 6 |
| 17 | 46–50 | M | SCA2 | 0.50 (1.22) | 3.00 | 0.60 | 0.17 (0.41) | 1 | 0.20 | 2 | 6 | 2.75 (3.49) | 8.00 | 4.29 | 0.75 (0.89) | 2 | 1.14 | 3 | 8 |

**Supplementary Table S3.** Finger Chase: agreement between the live-rated and video-derived SARA scores using the two methods of deriving the video-based score, as the SARA instructs (average of the last 3 of the first 5 trials) and from all trials. Matched = the live score equaled the video-derived score for that patient; mismatched = the two scores differed, and Live higher / Live lower give the direction (live score minus video score). 95% CI = Wilson score interval for the match rate.

|  | | **As instructed: average of the last 3 of the first 5 trials** | | | | | **All trials** | | | | |
| --- | --- | --- | --- | --- | --- | --- | --- | --- | --- | --- | --- |
| **Hand** | **Patients (N)** | **Matched, n (%)** | **95% CI** | **Mismatched, n (%)** | **Live score higher, n** | **Live score lower, n** | **Matched, n (%)** | **95% CI** | **Mismatched, n (%)** | **Live score higher, n** | **Live score lower, n** |
| Left | 17 | 11 (64.7%) | [41.3%, 82.7%] | 6 (35.3%) | 4 | 2 | 14 (82.4%) | [59.0%, 93.8%] | 3 (17.6%) | 2 | 1 |
| Right | 17 | 7 (41.2%) | [21.6%, 64.0%] | 10 (58.8%) | 4 | 6 | 12 (70.6%) | [46.9%, 86.7%] | 5 (29.4%) | 1 | 4 |
