## Supplementary Figures for "Challenges in Quantifying Upper Extremity Dysmetria in the Scale for the Assessment and Rating of Ataxia (SARA)"

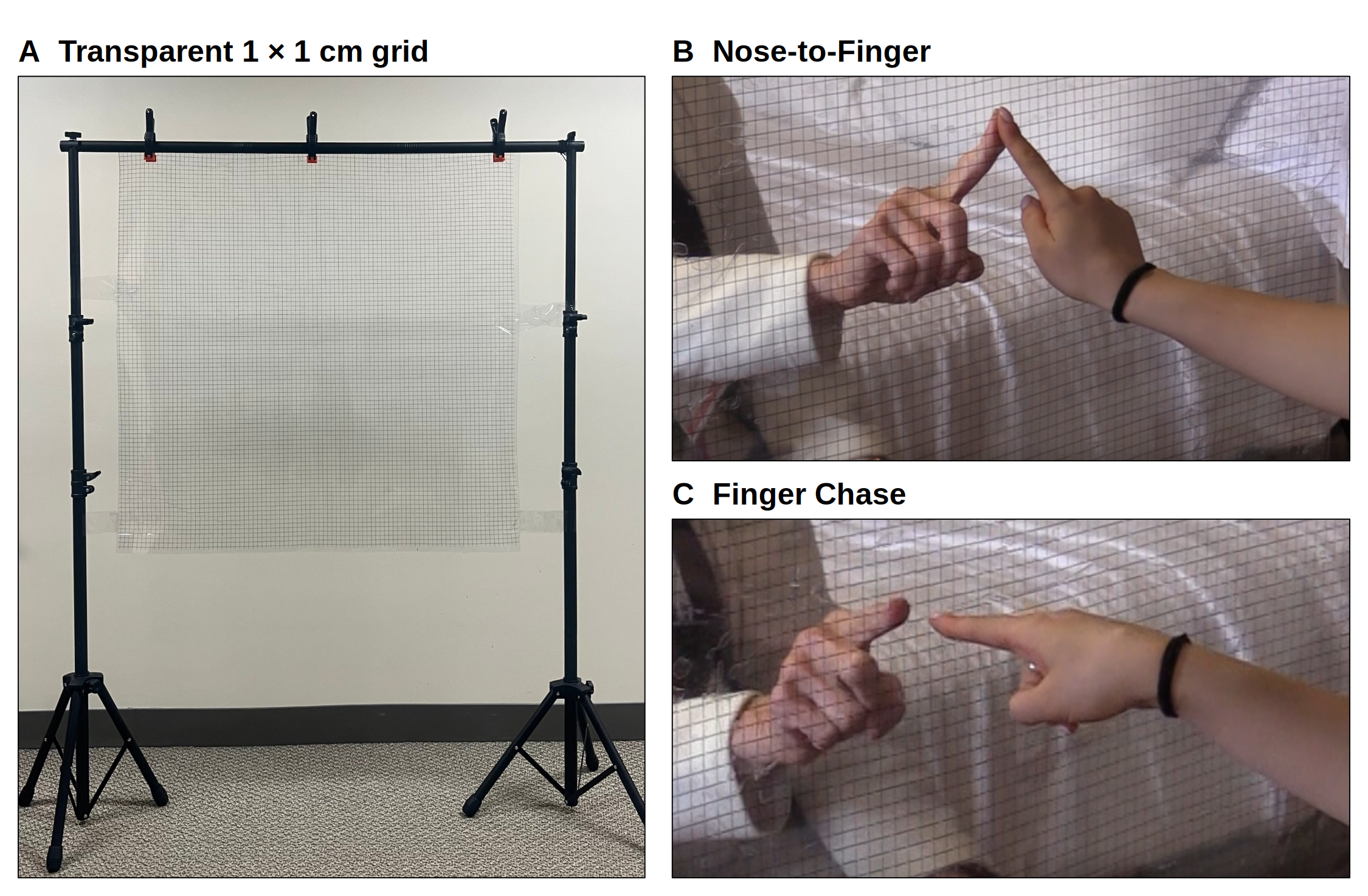

**Supplementary Figure S1. Testing Apparatus.** (A) Transparent 1 × 1 cm grid screen on its stand. (B) Nose-to-Finger: the subject touches the examiner’s finger on the screen. (C) Finger Chase: the subject follows the examiner’s finger, keeping their finger close to the screen without touching it.

**Supplementary Figure S2.**

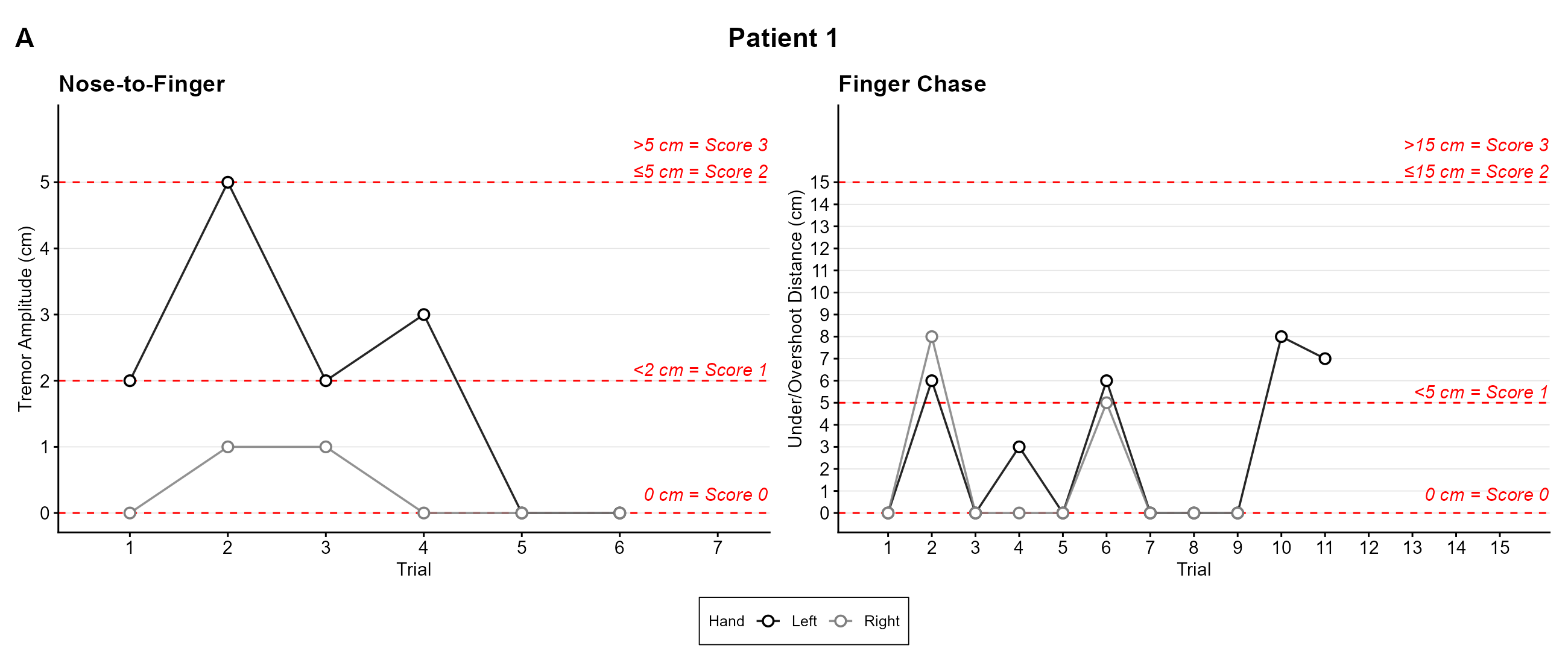

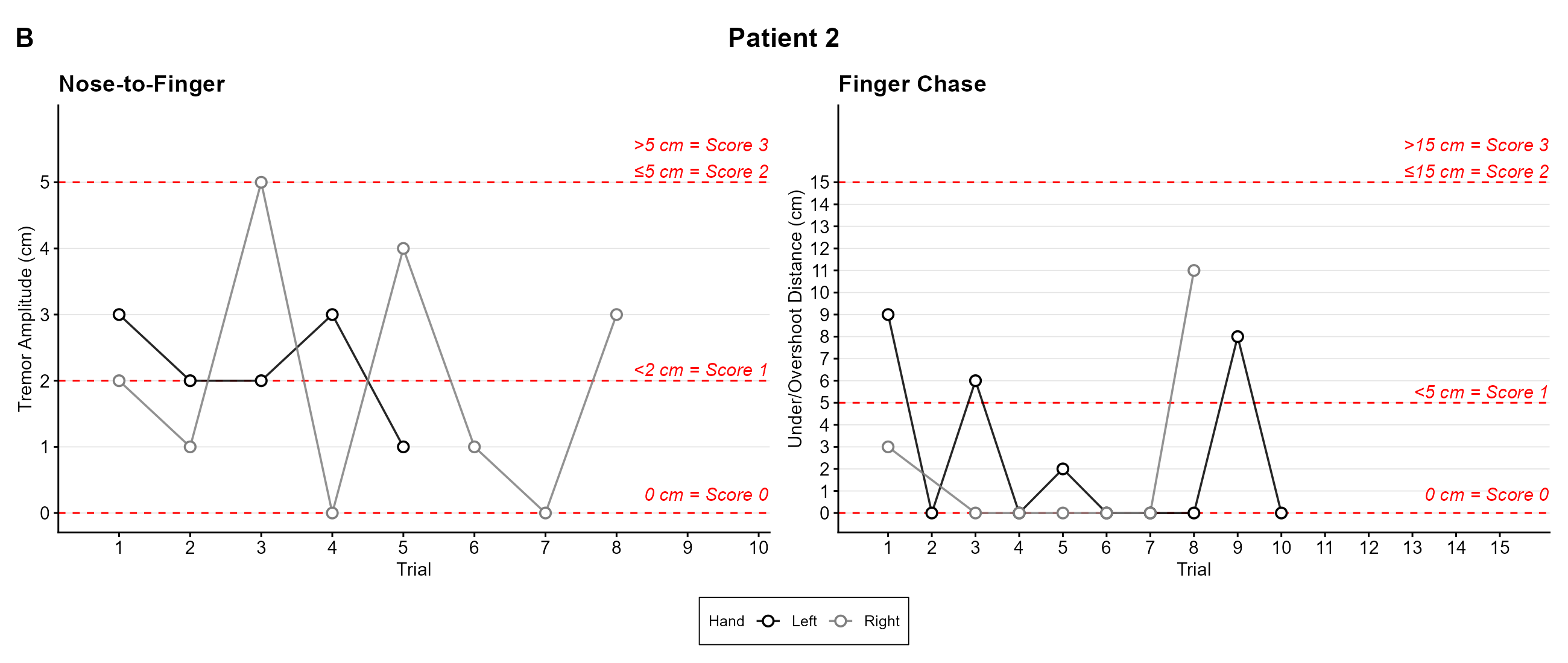

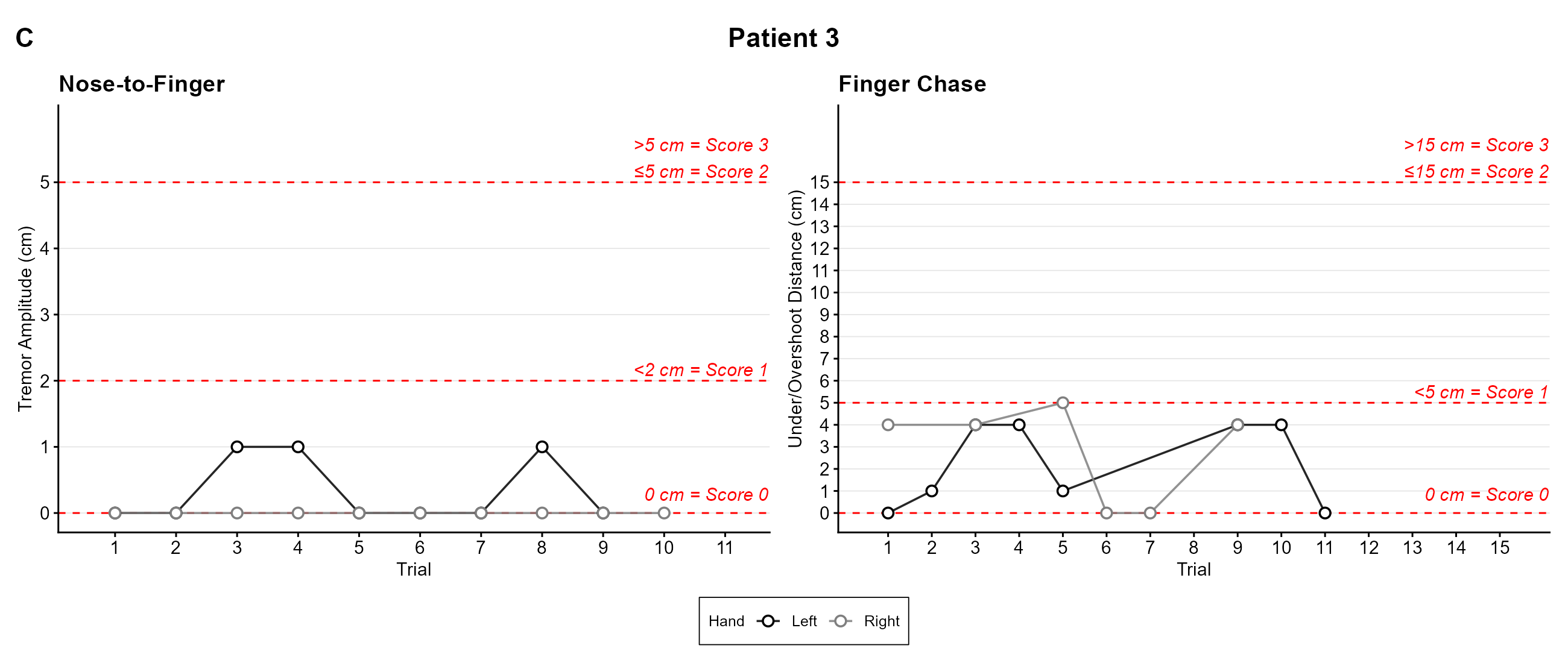

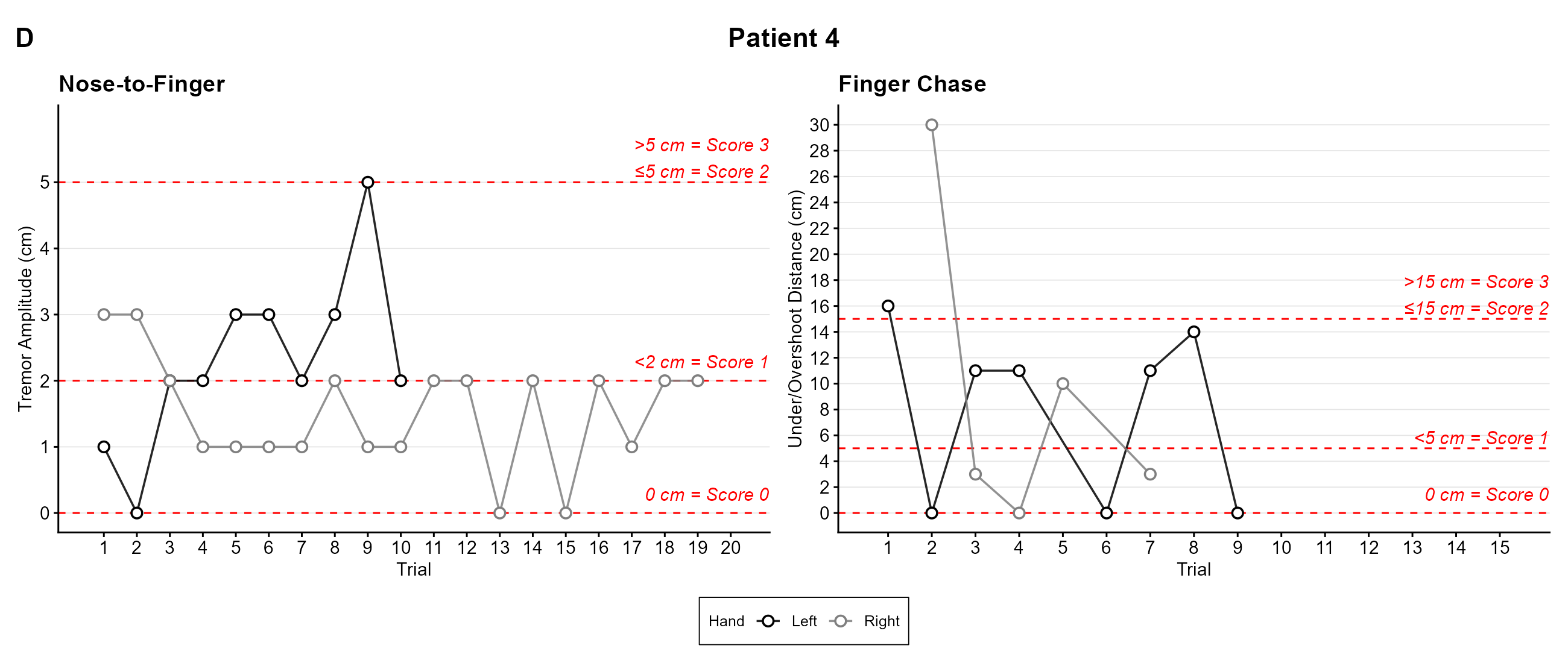

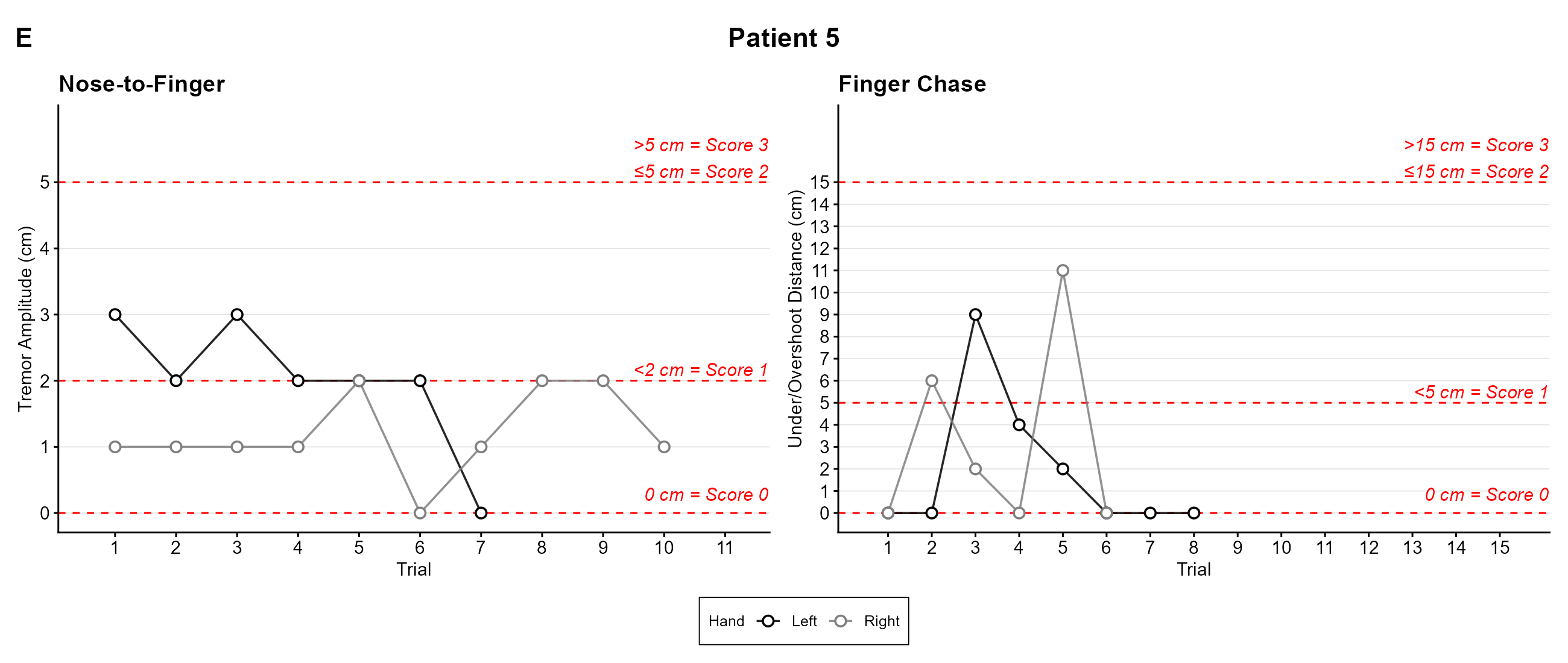

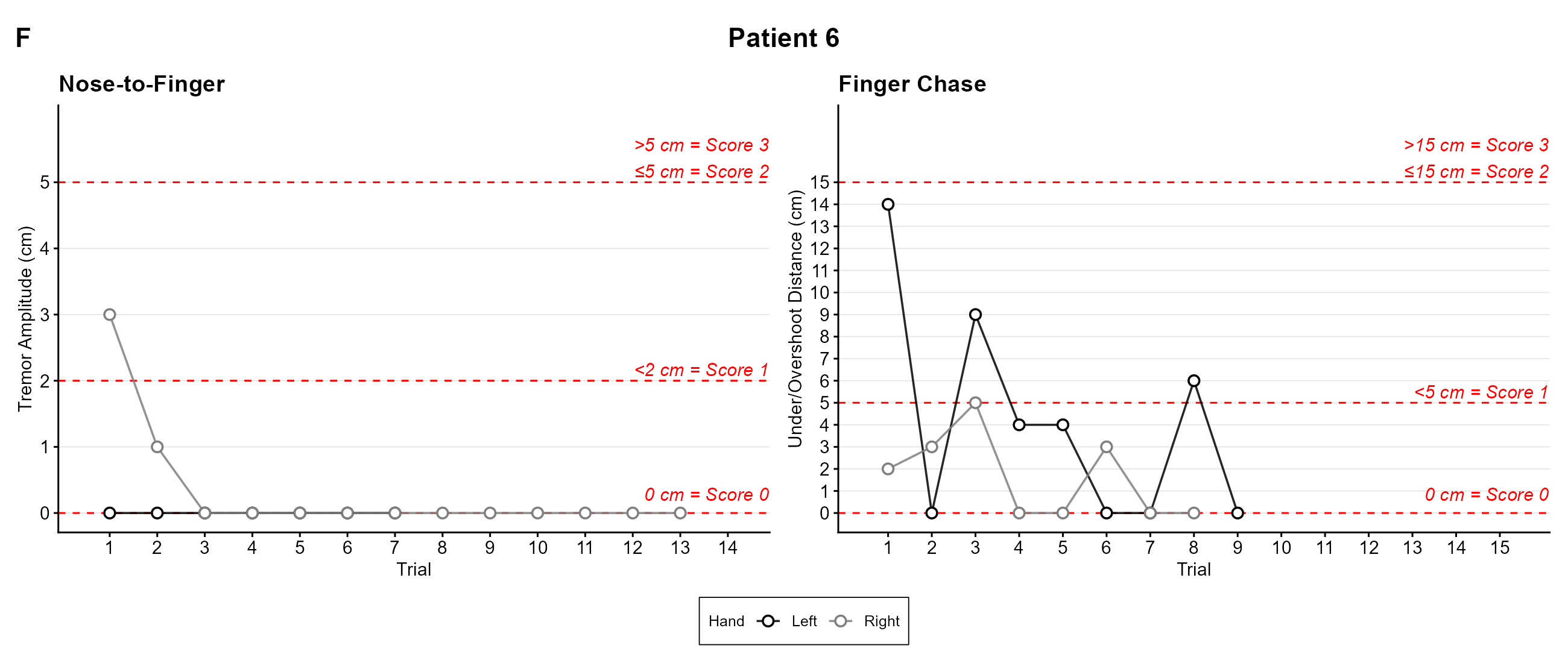

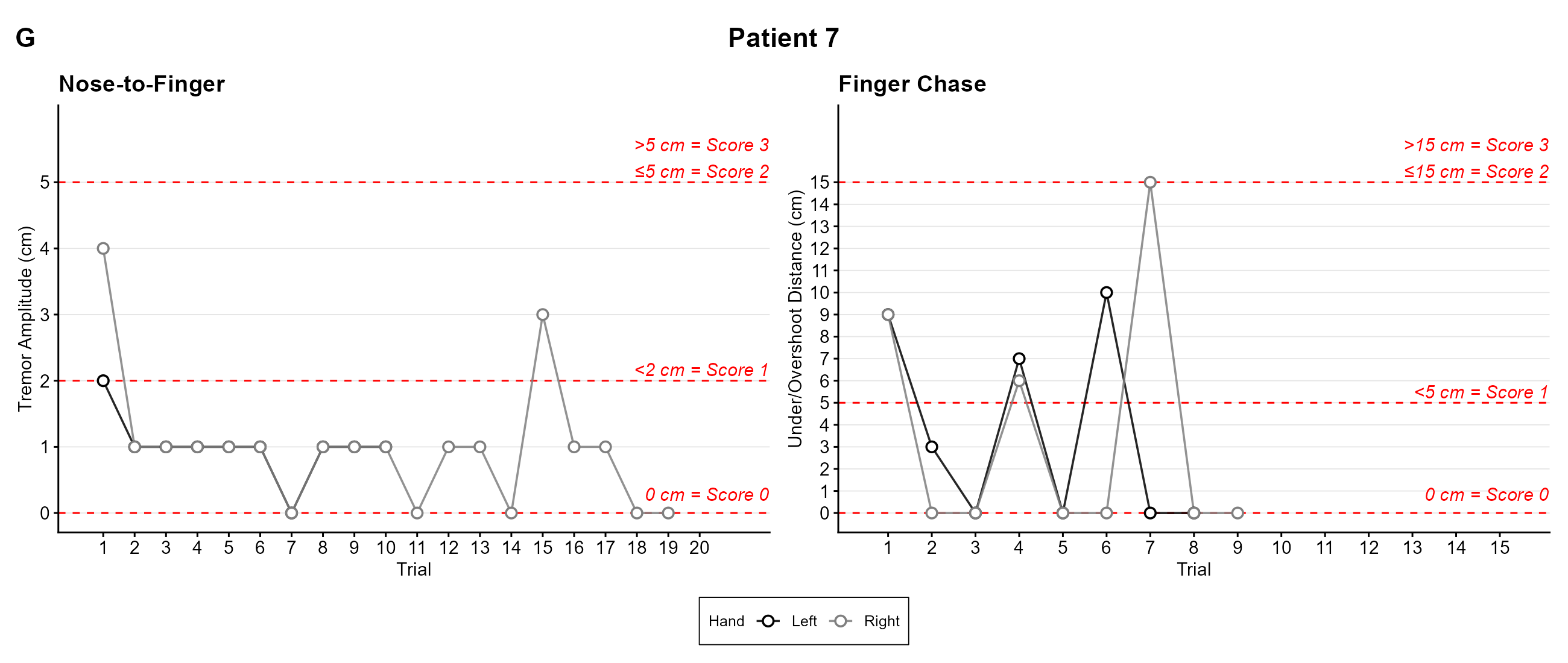

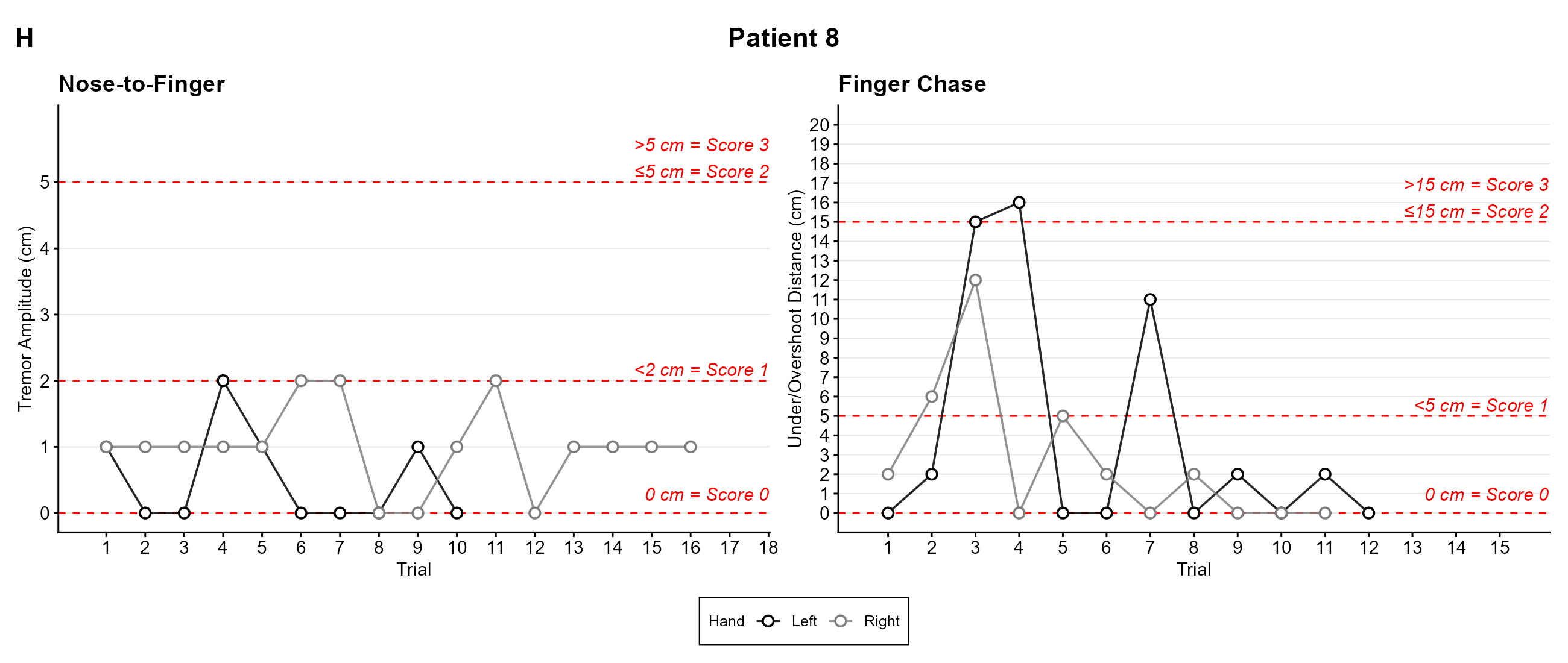

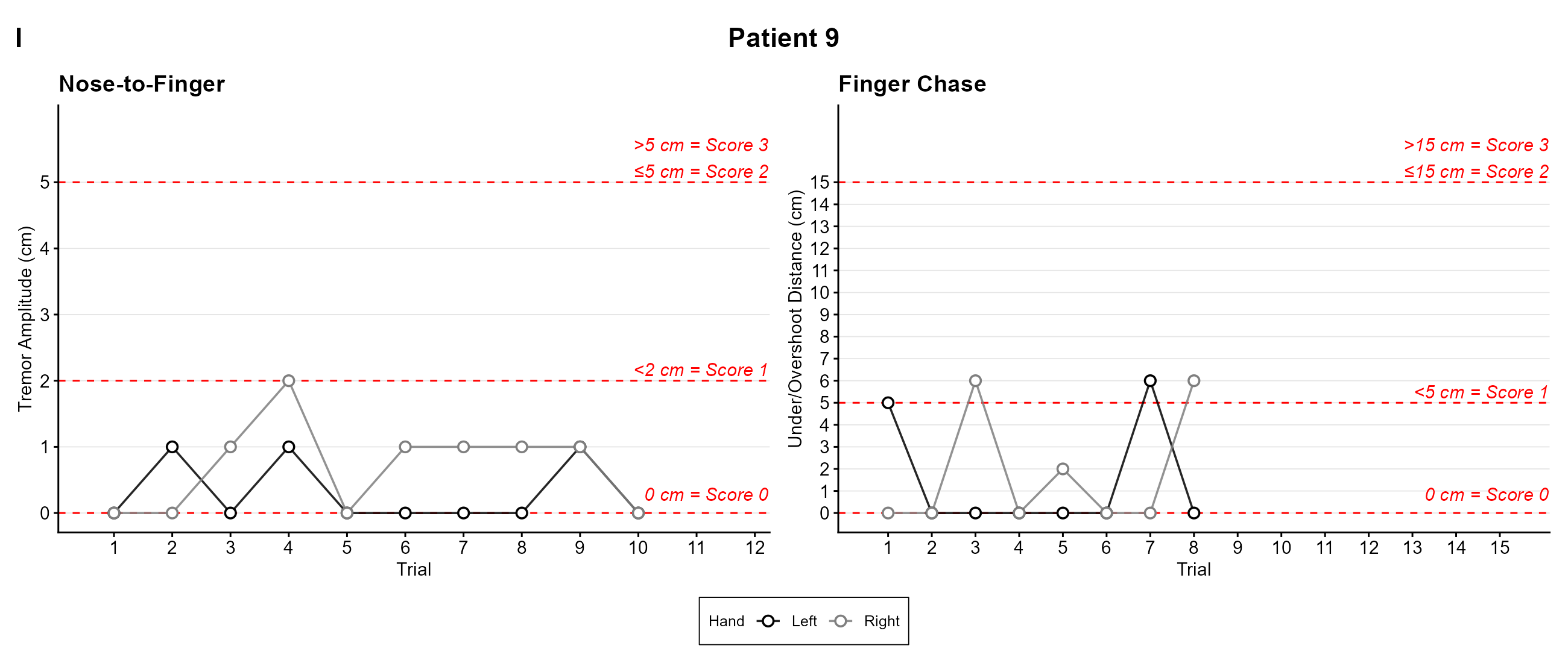

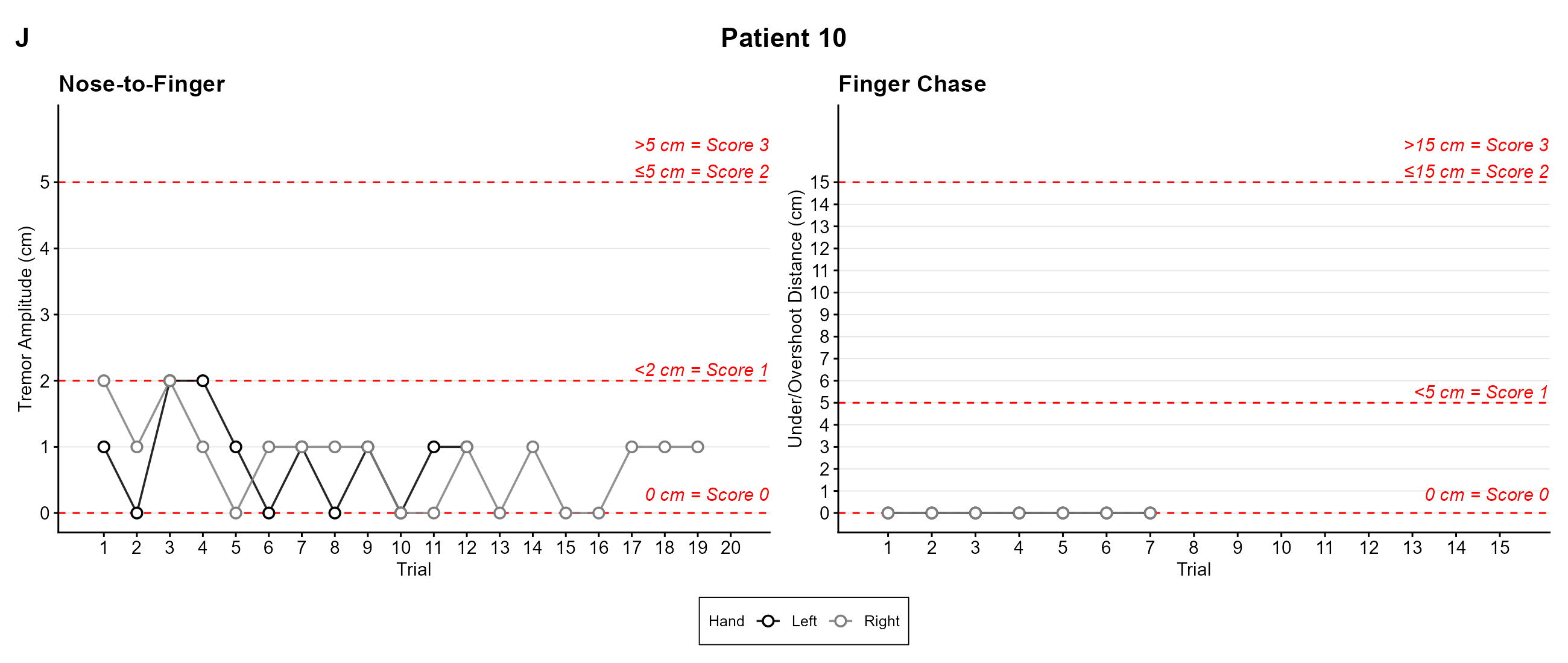

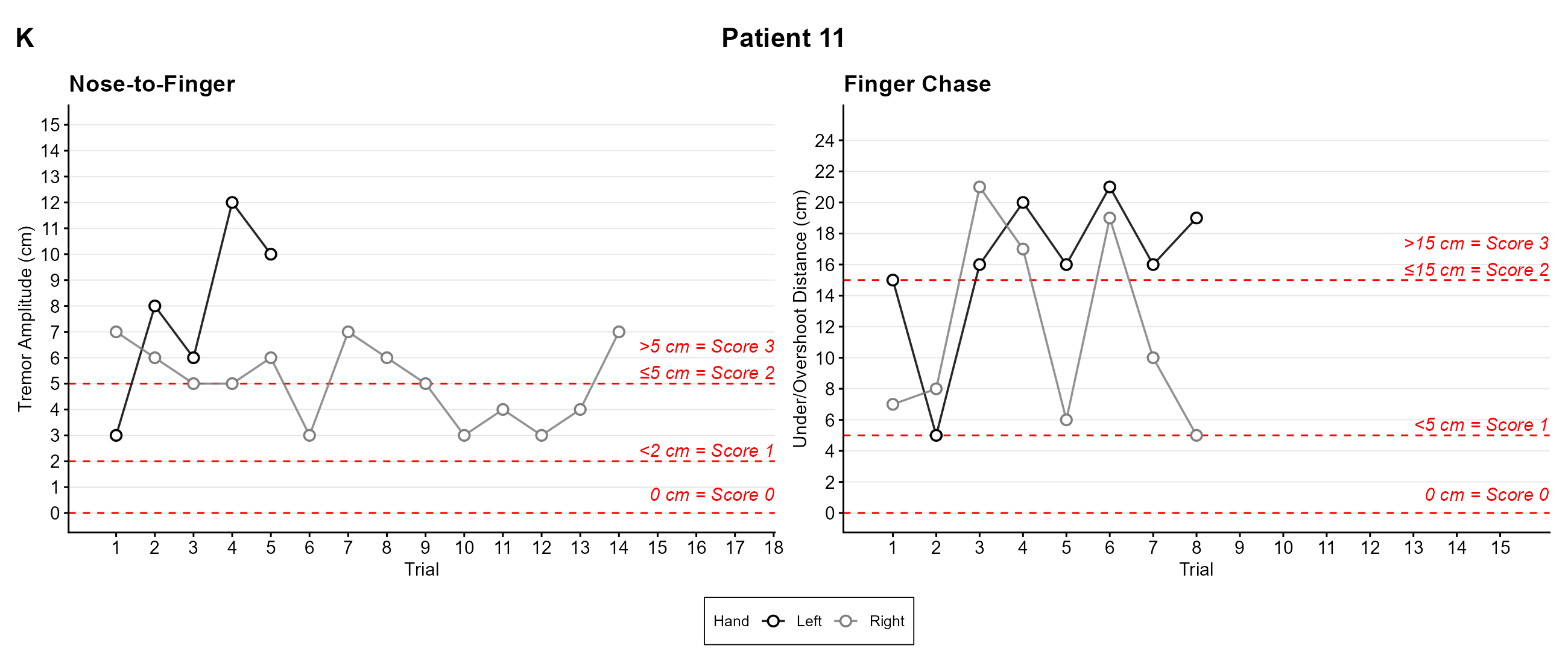

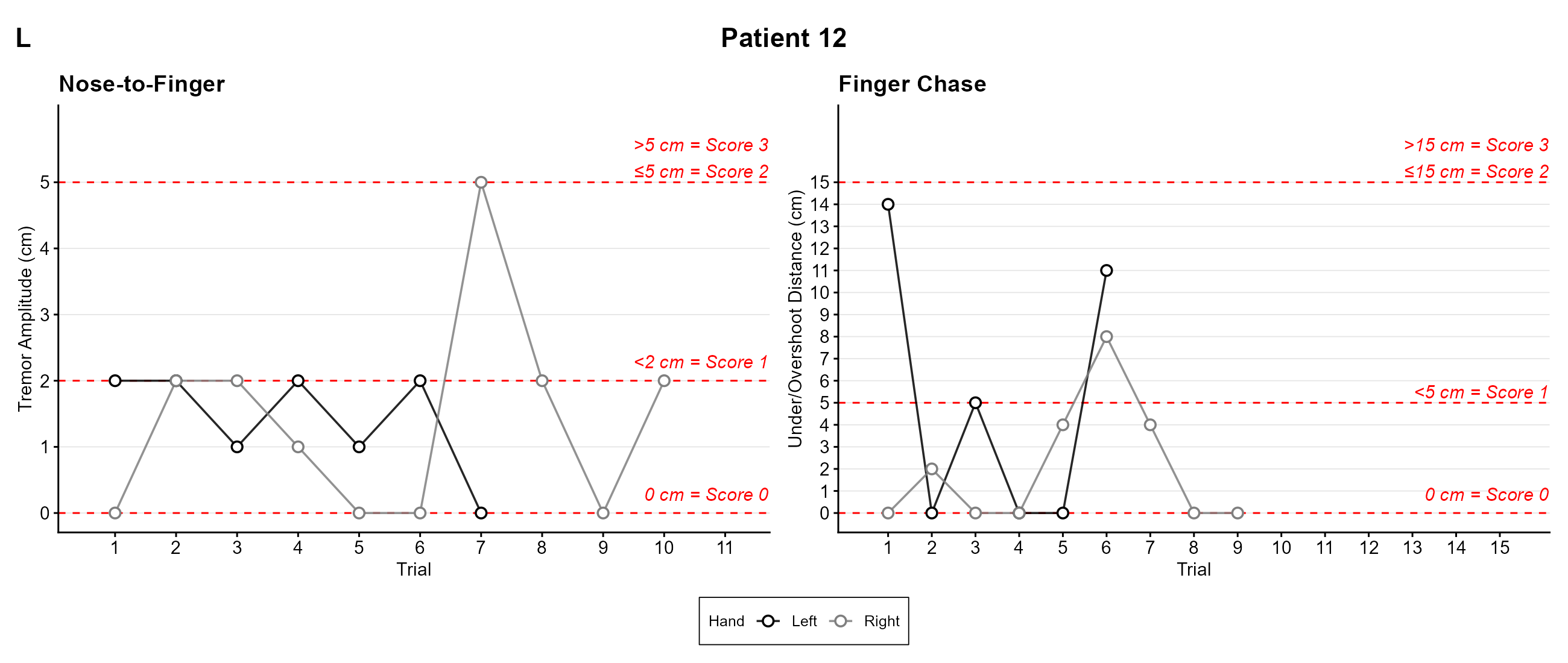

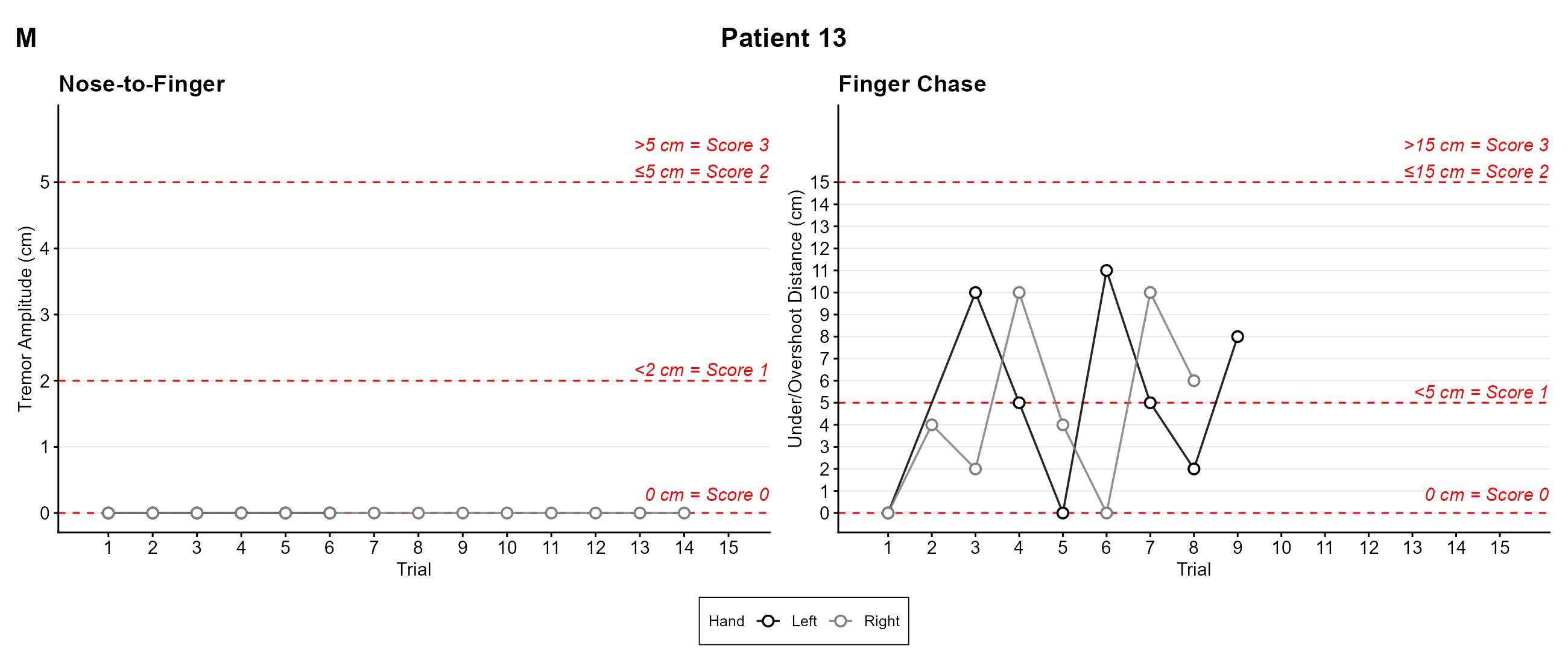

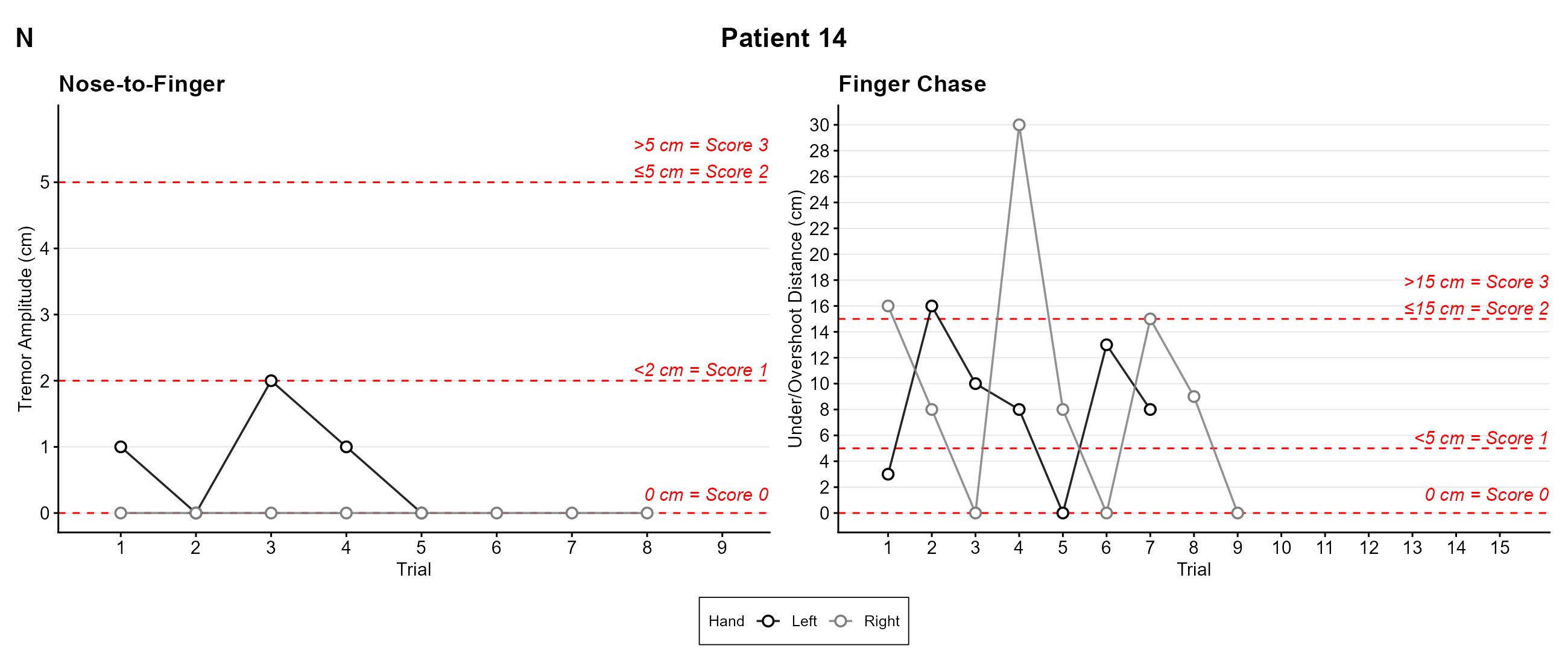

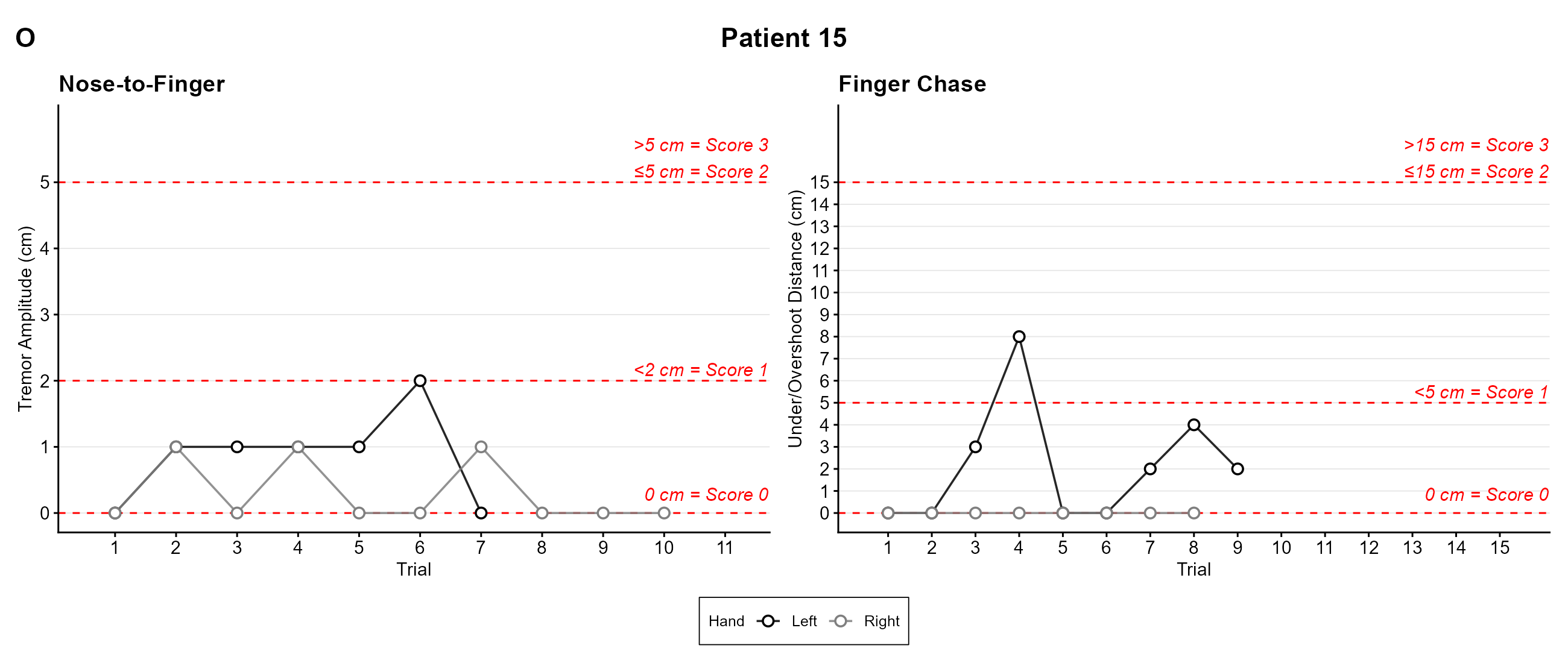

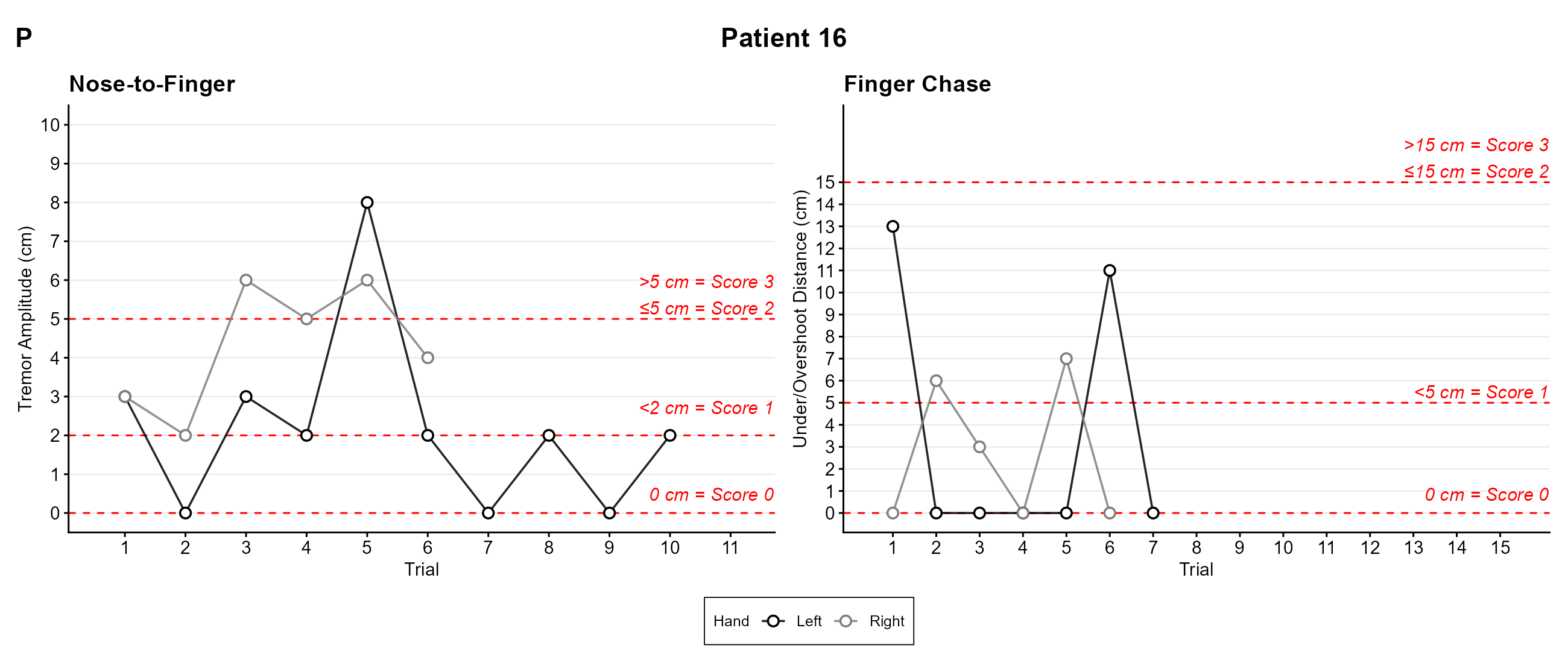

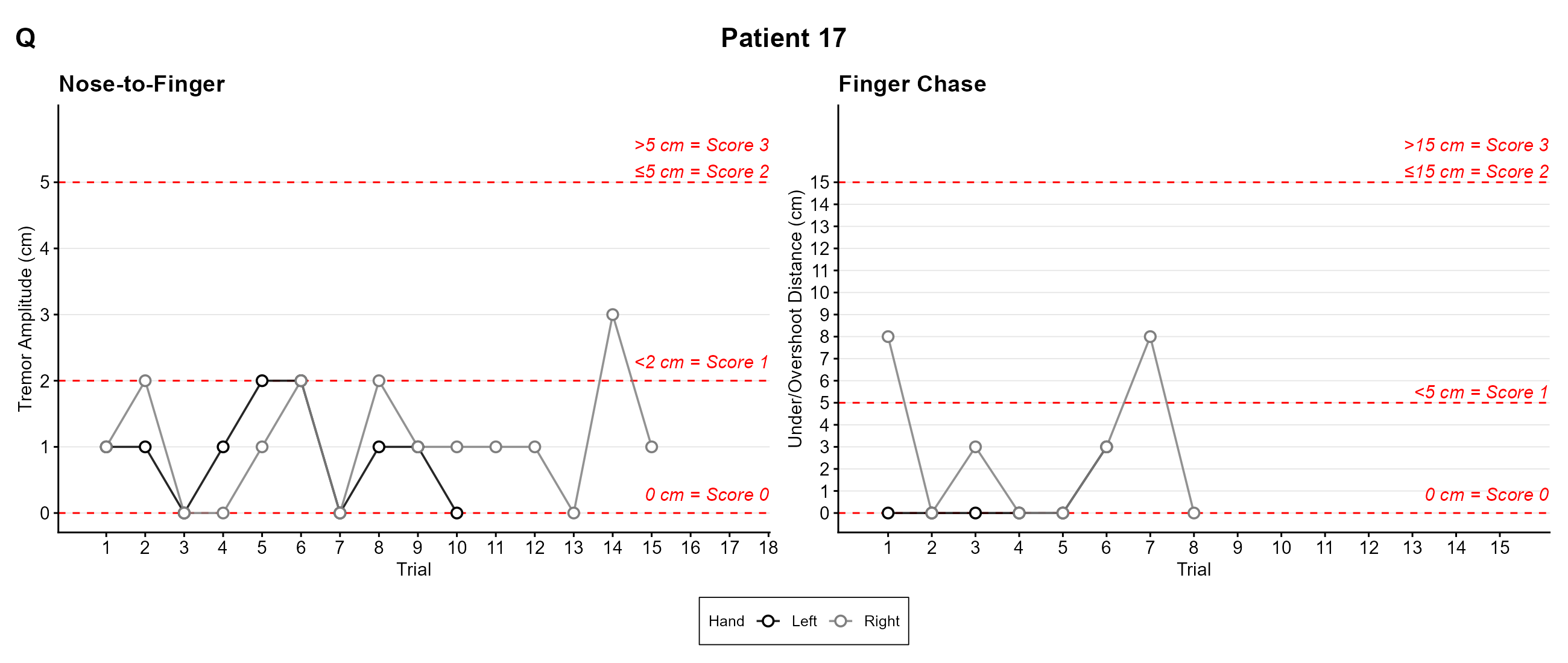

**Supplementary Figure S2.** Trial-by-trial measurements for each patient. Panels A–Q show patients 1–17. For each patient, the left plot shows nose-to-finger tremor amplitude (cm), and the right plot shows finger chase under/overshoot distance (cm) for every trial, for the left hand (black) and the right hand (grey). Red dashed lines mark the SARA score cut-offs.

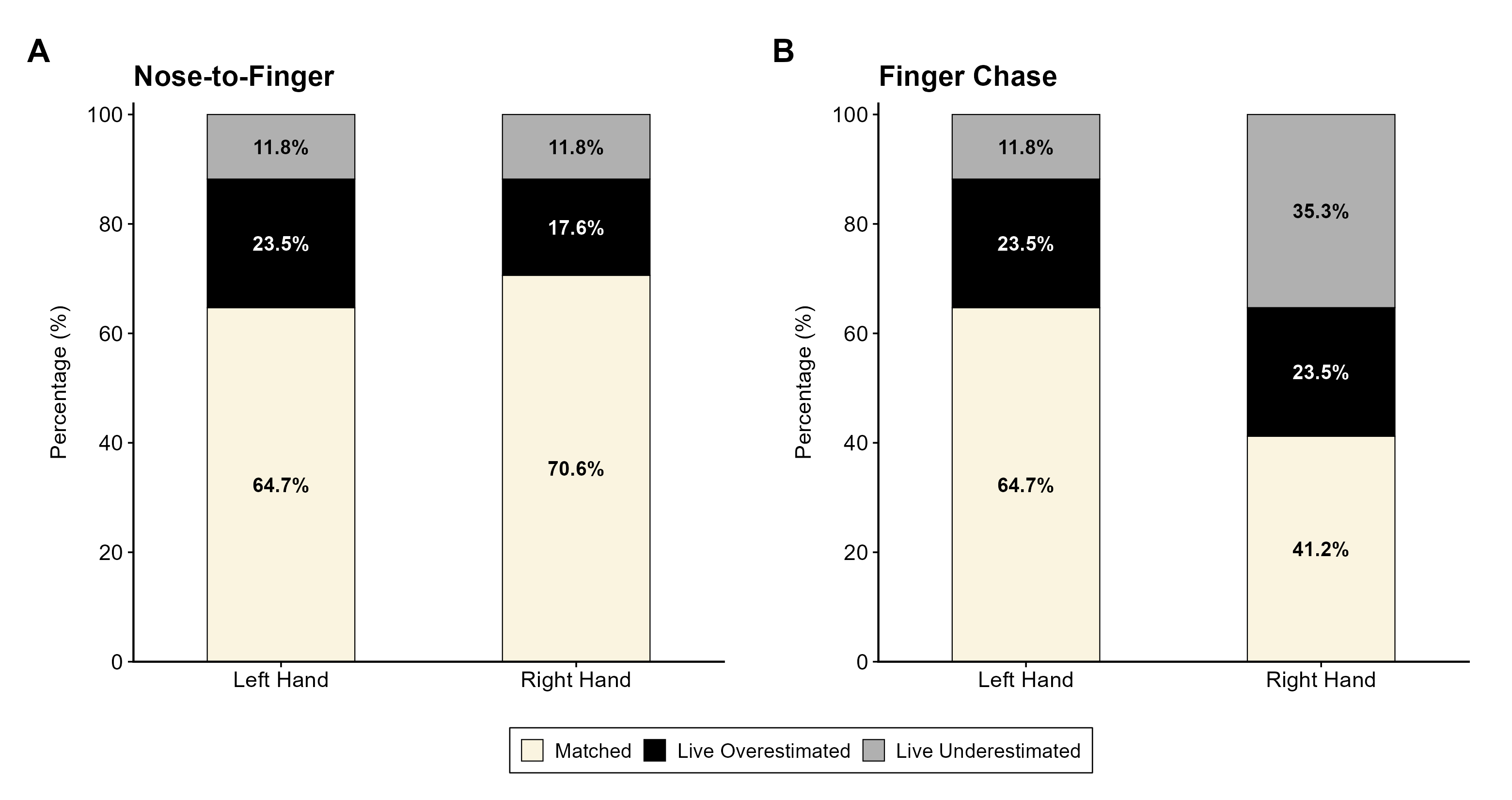

**Supplementary Figure S3.** Agreement between the live and video-derived SARA scores. Stacked bars show the percentage of patients (N = 17) for whom the live SARA score matched the video-derived score (beige), or the live score was higher than the video-derived score (live overestimated, black) or lower than the video-derived score (live underestimated, grey), for the left and right hands, in (A) SARA nose-to-finger and (B) SARA finger chase.
